# Confounder-adjusted MRI-based predictors of multiple sclerosis disability

**DOI:** 10.1101/2022.04.18.22273974

**Authors:** Yujin Kim, Mihael Varosanec, Peter Kosa, Bibiana Bielekova

## Abstract

**Introduction:** Both aging and multiple sclerosis (MS) cause central nervous system (CNS) atrophy. Excess brain atrophy in MS has been interpreted as accelerated aging. Current paper tests an alternative hypothesis: MS causes CNS atrophy by mechanism(s) different from physiological aging. Thus, subtracting effects of physiological confounders on CNS structures would isolate MS-specific effects.

**Methods:** Standardized brain MRI and neurological examination were acquired prospectively in 649 participants enrolled in ClinicalTrials.gov Identifier: NCT00794352 protocol. CNS volumes were measured retrospectively, by automated Lesion-TOADS algorithm and by Spinal Cord Toolbox, in a blinded fashion. Physiological confounders identified in 80 healthy volunteers were regressed out by stepwise multiple linear regression. MS specificity of confounder-adjusted MRI features was assessed in non-MS cohort (n=160). MS patients were randomly split into training (n=277) and validation (n=132) cohorts. Gradient boosting machine (GBM) models were generated in MS training cohort from unadjusted and confounder-adjusted CNS volumes against four disability scales.

**Results:** Confounder adjustment highlighted MS-specific progressive loss of CNS white matter. GBM model performance decreased substantially from training to cross-validation, to independent validation cohorts, but all models predicted cognitive and physical disability with low p-values and effect sizes that outperforms published literature based on recent meta-analysis. Models built from confounder-adjusted MRI predictors outperformed models from unadjusted predictors in the validation cohort.

**Conclusion:** GBM models from confounder-adjusted volumetric MRI features reflect MS-specific CNS injury, and due to stronger correlation with clinical outcomes compared to brain atrophy these models should be explored in future MS clinical trials.

**Highlights:**

1. Regressing out physiological confounders affecting volume of CNS structures in healthy volunteers, strengthened correlations between white matter volumes and disability outcomes in MS cohorts
2. Aggregating volumetric features into generalized boosting machine (GBM) models outperformed correlations of individual MRI biomarkers with clinical outcomes in MS
3. Developed more sensitive and reliable models that predict MS-associated disability
4. Independent validation cohorts show true model performances
5. Developed GBM models should be explored in future MS clinical trials

## 1. INTRODUCTION

Scientific advancements in Multiple Sclerosis (MS) translated into the development of disease-modifying treatments (DMTs) that effectively inhibit central nervous system (CNS) tissue destruction and clinical disability development if given to a young subject shortly after disease onset. Unfortunately, these treatments lose efficacy with advancing age of patients. On a group level, they show no statistically-significant inhibition of disability progression in subjects older than 54 years (Weideman et al., 2017b). To develop more effective MS treatments, we need sensitive outcomes that can objectively quantify CNS tissue destruction and development of disability against active comparator, in reasonably sized cohorts.

Magnetic resonance imaging (MRI) volumetric outcomes such as brain atrophy, have been successfully used in Phase II and Phase III MS clinical trials (Hemond and Bakshi, 2018; Jacobs et al., 2021; Zeydan and Kantarci, 2020). However, is brain atrophy the most sensitive and the most specific imaging outcome in MS?

In terms of sensitivity, there is mounting evidence that atrophy of deep gray matter (GM) structures, especially thalamus (Amin and Ontaneda, 2021), or enlargement of ventricles (Jakimovski et al., 2020) may exert larger effect sizes in MS than whole brain atrophy. Additionally, assembling several of these changing brain volumes into a single model using machine-learning (ML) algorithm(s) may further increase effect size.

In terms of specificity, brain atrophy also occurs during natural aging (Rocca et al., 2017). In what we identified as the largest published study of healthy volunteers (HV; n=2790), physiological confounders such as age, gender and predicted intracranial volume (PIV), explained a very high proportion of variance in thalamic (>60%), caudate (>40%) and ventricular (57%) volumes (Potvin et al., 2016). In fact, ML-derived models can predict chronological age (with a mean absolute error of +/-5 years (Cole et al., 2017)) using just brain MRI predictors. A study from European Magnetic Resonance Imaging in MS (MAGNIMS) consortium (Cole et al., 2020) explored the difference between chronological and brain MRI-predicted age, termed as Brain-predicted Age Difference (Brain-PAD). The study demonstrated that 1204 MS patients had on average 6 years higher Brain-PAD compared to HV, without significant differences among MS subtypes (i.e., relapsing-remitting [RRMS], secondary-[SPMS] and primary-progressive MS [PPMS]). Brain-PAD also correlated with disability measured by Expanded Disability Status Scale (EDSS). Although no effect size was provided, the associated figures suggested that Brain-PAD explains < 10% of EDSS variance. The authors concluded that MS is associated with accelerated brain aging (Cole et al., 2020).

It could be alternatively interpreted that MS and natural aging destroy structurally overlapping CNS areas, but MS does so by different pathophysiological processes. This interpretation may explain why treated MS patients had significantly higher Brain-PAD compared to untreated MS patients (Cole et al., 2020). If MS causes accelerated brain aging and MS DMTs inhibit MS-associated CNS damage, then treated MS patients should have decreased Brain-PAD, not increased. On the other hand, if MS and natural aging cause CNS damage by different mechanisms, then MS DMTs may have none, or even paradoxical effect(s) on Brain-PAD. Our previous study demonstrated that CSF biomarkers also predict chronological age in HV with high accuracy. When we applied this model to MS patients, we saw no significant differences in age-predictions between HV and MS (Barbour et al., 2020). These data strongly support this alternative interpretation, in which (molecular) mechanisms of aging and MS progression are mostly different.

Further extension of this interpretation is the hypothesis that aging process and other physiological confounders reflected on volumetric brain MRI represent “noise” when trying to understand (and treat) MS-specific processes. Here, we examine this hypothesis by addressing the following aims: (1) To adjust volumes of the CNS structures for physiological confounders identified from internal HV cohort to understand effects of MS on CNS structures; (2) To examine, whether confounder-adjusted MRI predictors can be assembled into computational models that predict clinical disability in the independent validation cohort, and whether such validated model exerts larger effect size(s) than any single MRI biomarker; (3) To investigate whether computational model(s) derived from confounder-adjusted MRI predictors outperform models(s) from raw MRI volumes in predicting clinical disability outcomes.

## 2. MATERIAL AND METHODS

The study design is shown in Figure 1.

**Figure 1.**
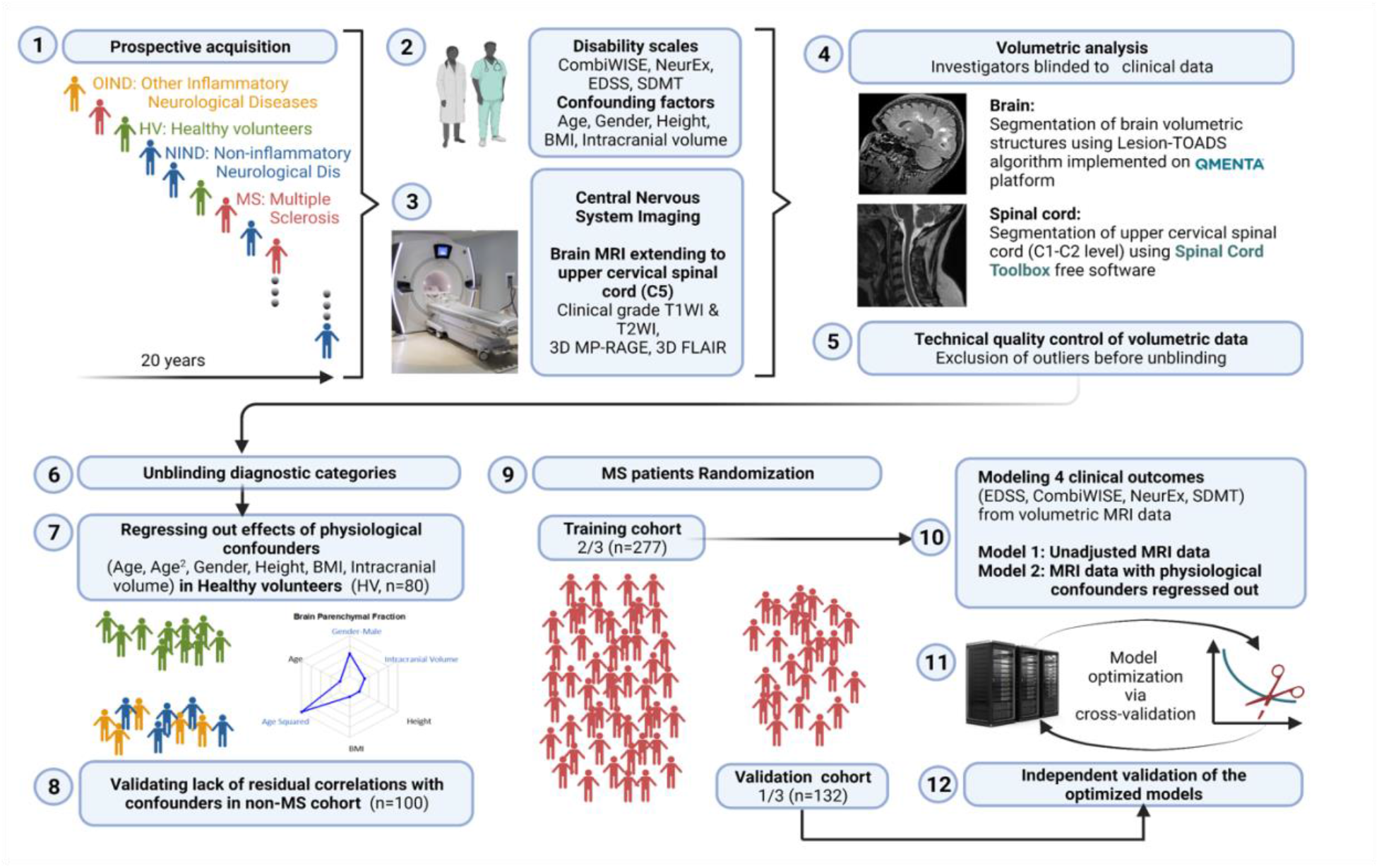
Study design. **1:** All subjects participating in a prospective collection of standardized clinical and imaging outcomes under a natural history protocol for 20 years were enrolled. **2:** All subjects underwent full neurological examination transcribed to the NeurEx™ App that automatically calculates clinician-derived disability scales. Additional functional tests, such as Symbol Digit Modalities Test (SDMT) and stated confounding factors were collected and transcribed to research database. **3:** All subjects underwent research brain MRI that extended caudally to the C5 level of the spinal cord (SC). **4:** Anonymized MRIs were uploaded to the cloud-based QMENTA platform to derive brain volumetric data using Lesion-TOADS algorithm. Upper cervical SC C1-C2 volume was calculated using Spinal Cord Toolbox. **5:** Resulting quantitative MRI biomarkers were assessed for quality to identify intra- and inter-individual outliers. Identified outliers were manually checked and scans with incorrect segmentation were excluded (122/771 = 15.8%). **6:** Unblinding of diagnostic categories occurred after exclusion of technical outliers was completed. **7:** We regressed out the effects of six stated confounders measured in the HV cohort (n=80) and applied the same transformation to non-MS (n=100) and MS (n=409) cohorts to eliminate effects of physiological confounders on MRI volumes. **8:** We validated lack of residual correlations with the confounding factors in non-MS cohort. **9:** MS patients were randomly split into training (n=277) and validation (n=132) cohorts using stratified split to assure equal proportions of gender and MS types in both cohorts. **10:** Gradient boosting machine (GBM) algorithm was applied to the training cohort using confounder-adjusted (and unadjusted) MRI features as predictors to derive two sets of models for the four stated clinical outcomes. **11:** Models were further optimized in the training cohort using 10-fold cross validation. **12:** Resulting eight models were applied to the independent validation cohort that did not contribute, in any way, to the generation or optimization of the models.

### 2.1 Cohort characteristics

All patients were prospectively recruited into the protocol “Comprehensive Multimodal Analysis of Neuroimmunological Diseases of the Central Nervous System” (Clinicaltrials.gov Identifier: NCT00794352) and provided written informed consent. The inclusion criteria for patient cohort are age at least 12 years and clinical symptoms, CSF results or MRI imaging suggestive of neuroimmunological disease. Approximately 60% of enrolled patients eventually fulfill contemporary version of MS diagnostic criteria (with all 3 MS subtypes represented), while 20% have other inflammatory neurological diseases (OIND) and 20% have non-inflammatory neurological diseases (NIND). The HV inclusion criteria are age at least 18 years, absence of known diseases and conditions that could affect CNS and normal vital signs at the screening visit. The protocol was approved by the Combined Neuroscience Institutional Review Board of the National Institutes of Health. Patient demographics and other clinical characteristics are provided in Supplementary Figure S1.

All subjects (n=649) with brain MRI images that passed quality control (see below) and had matched clinical outcomes were included. Neurological exams documented in structured electronic medical record note were either transcribed (before 2017) or directly documented (after 2017) into the NeurEx™ App (Kosa et al., 2018) by clinicians with MS specialization. The NeurEx™ App automatically computes MS disability scales including EDSS (Kurtzke, 1983) (ordinal scale from 0-10) and NeurEx (continuous scale from 0 to theoretical maximum of 1349).

Functional tests (i.e., timed 25 foot walk and 9 hole peg test) are required for computing of Combinatorial weight-adjusted disability score (CombiWISE; continuous scale from 0-100) (Weideman et al., 2017a), and Symbol Digit Modalities Test (SDMT) (Benedict et al., 2017). These were acquired by investigators blinded to clinician-derived disability scales and were recorded into research database.

### 2.2 Volumetric Analysis

Investigators involved in MRI analysis were blinded to clinical outcomes and diagnostic categories. MRIs were performed on two scanners: Signa (3TA, General Electric, Milwaukee, WI) using 16-channel head coil and Skyra (3TD, Siemens, Malvern, PA) using 32-channel head coil. Sequences included 3D-MPRAGE (TR, 3000 ms; TE, 3 ms; TI, 900 ms; FA 8°; 1-mm isotropic resolution, TA 6 min), 3D-FLAIR (TR, 4800 ms; TE, 354 ms; TI, 1800 ms; 1-mm isotropic resolution; acquisition time, 7 min), and PD/T2 (TR, 3540 ms; TE, 13 and 90 ms; 0.8-mm in-plane resolution; slice thickness, 2 mm; acquisition time, 4 min) on 3TD and 3D-FSPGR-BRAVO (TR, 1760 ms; TE, 3 ms; TI, 450 ms; FA 13°; 1-mm isotropic resolution; acquisition time, 5 min), 3D-FLAIR-CUBE (TR, 6000 ms; TE, 154 ms; TI, 1800 ms; 1-mm isotropic resolution; acquisition time, 9 min), and PD/T2 (TR, 5325 ms; TE, 20 and 120 ms; 1-mm in-plane resolution; slice thickness, 3 mm; acquisition time 4 min) on 3TA. Sagittal and axial cuts extended distally to C5 level, allowing determination of semi-qMRI biomarkers of medulla/upper spinal cord (SC) atrophy using SC Toolbox (De Leener et al., 2017).

After pre-processing steps (1. De-identification, 2. DICOM to NIFTI transformation, 3. Skull stripping; and 4. Alignment), images were uploaded to commercial cloud-based imaging platform QMENTA (https://www.qmenta.com/) which, in collaboration, implemented the published Lesion-TOADS (Shiee et al., 2010) algorithm. Lesion-TOADS combines a topological and statistical likelihood atlas for computation of 12 CNS volumetric biomarkers: Cerebral white matter (WM), Cerebellar WM, Brainstem, Putamen, Thalamus, Caudate, Cortical gray matter (GM), Cerebellar GM, Lesion Volume, Ventricular CSF and Sulcal CSF. Average cross-sectional area (CSA) of the upper cervical SC at C1-C2 level was calculated using SC Toolbox from 3D MPRAGE brain MRI images.

Before unblinding, we performed quality control of volumetric data. A total of 62 (8.7%) MRI scans was excluded due to inaccurate segmentation of brain structures, low image quality, and motion artifacts, leaving 649 scans for the final dataset in study.

After unblinding diagnostic categories, MS patients were randomly split into training and validation cohort (2:1 ratio) with equal proportion in gender and MS subtypes.

### 2.3 Adjusting MRI biomarkers for healthy volunteers (HV) confounders

Using only the HV cohort (n=80), MRI volumes were adjusted for age, age^2, body mass index (BMI), height, gender, and supratentorial intracranial volume using stepwise multiple linear regression. Final linear regression models were applied to all subjects to regress out physiological confounders. The example of confounder adjustments for ventricular volume is shown in Figure 2. Supplementary information contains analogous data for all remaining MRI biomarkers.

**Figure 2.**
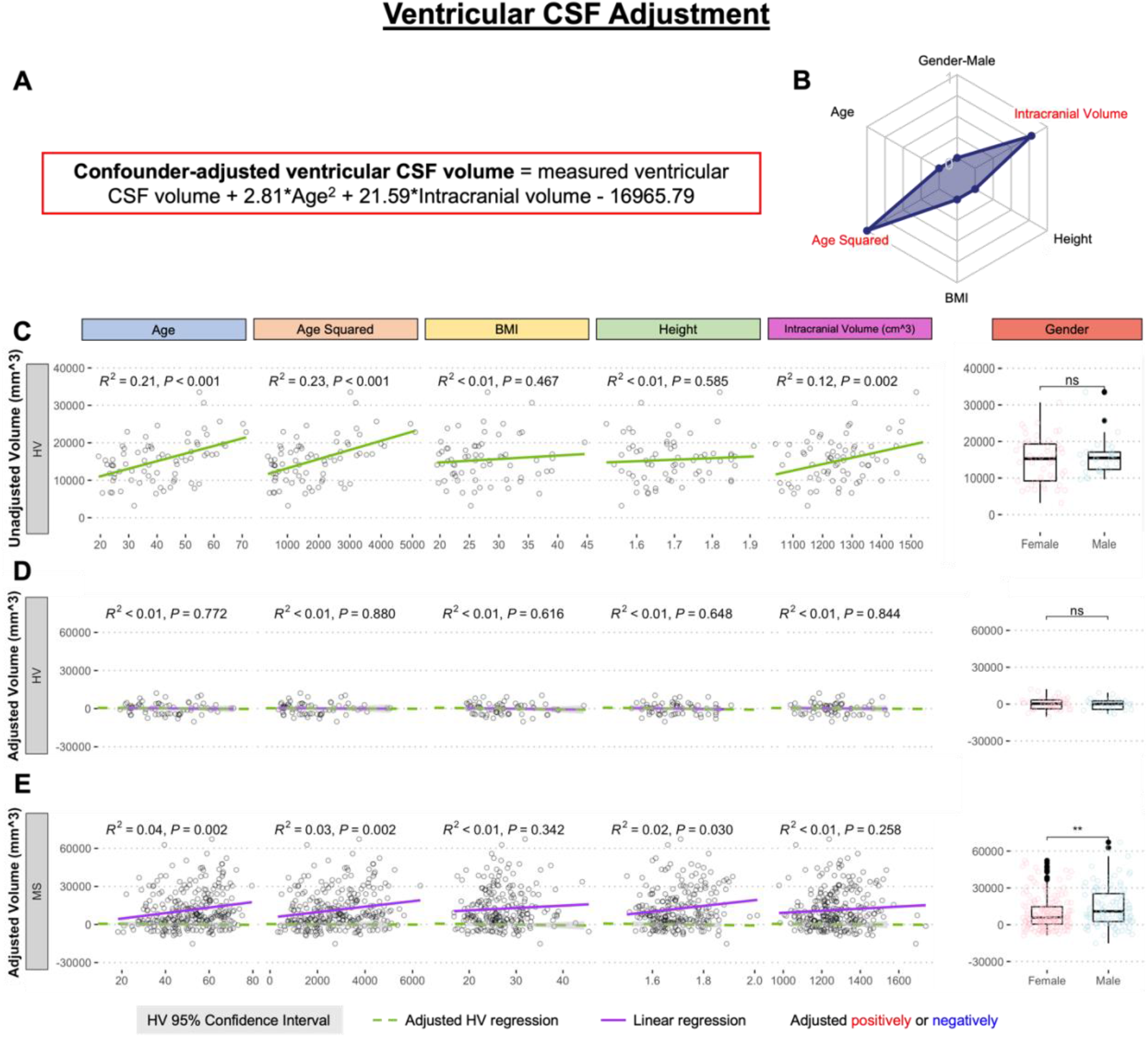
Example of the adjustment of single MRI biomarker (i.e., ventricular CSF volume) for six physiological confounders. **A**. The final equation for the confounder-adjusted ventricular volume is shown at the top of the Figure in red outline. This equation was derived from multiple linear regression models as described in the Method section. **B**. In the top right corner is resulting radar chart that show proportional weights of the applied confounder adjustment, with confounders with lowest weights (i.e., the innermost circle) representing zeros. **C**. Univariate linear regression models between each tested confounder on x-axis (first 5 graphs) or gender (sixth graph) and measured ventricular volume on the y-axis in 80 healthy volunteers (HV). **D**. Same univariate regressions in the HV cohort after applying adjustment formula show no remaining effect of confounders. **E**. Applying HV-derived adjustment formula to MS cohort shows remaining significant residual effect of age, when considering Bonferroni adjustment for multiple comparisons (i.e., p<0.05/12 = p<0.004). Analogous Figures showing adjustment for all MRI features are in the Supplementary information.

The correlation between the confounders and individual structural volumes, before and after co-variate adjustments, was evaluated using linear regression models in R Studio, reporting a coefficient of determination (R^2; the proportion of variance explained [ranging from 0 to 1] by the linear model, where higher number signifies closer fit of experimental data points to the linear regression line of the model), slope, and HV 95% confidence interval.

Differences between HV and MS cohorts in unadjusted and confounder-adjusted MRI biomarkers were analyzed using unpaired two-samples Wilcoxon signed-rank test in R (Team, 1969).

### 2.4 Gradient Boosting Machine (GBM) Modeling in the MS training cohort

Unadjusted or confounder-adjusted MRI biomarkers that showed statistically significant difference between MS and HV in univariate analyses were used as predictors to model four clinical outcomes: CombiWISE, EDSS, NeurEx, and SDMT. We selected a tree-based supervised ML algorithm because we considered that MRI features may have non-linear effects and patients may exert heterogeneity in which brain/SC structures are most affected by the disease. Among tree-based algorithms, we selected GBM over Random Forest. While GBM is more difficult to optimize (if using large number of predictors), it is believed to generally outperform Random Forest. GBM builds trees sequentially, where each successive tree is built using residuals from the previous tree’s predictions. The predictions are iteratively updated by adding the current tree’s prediction (times a shrinkage parameter) to the previous tree’s prediction. For each tree constructed, an out-of-bag (OOB) sample containing half of the observations is withheld from the training cohort to introduce randomness into the modeling process. Main GBM tuning parameters are the depth of the individual trees (interaction depth), the shrinkage parameter (learning rate), the minimum number of observations in trees’ terminal nodes, and the number of trees. Using the *gbm* R package (Greenwell et al., 2020), we selected an interaction depth of 6, nodes of 5, a shrinkage parameter of 0.01, and used a 10-fold cross validation to select the optimal number of trees that will prevent each model from overfitting.Improvement in mean squared error from splits within each individual tree and the average of these improvements across all trees in the ensemble calculated the relative influence of each variable in a model.

Each clinical scale was modelled separately since each assesses different attributes of the neurological spectrum (e.g., SDMT measures reaction time reflective of cognitive disability whereas other three scales reflect predominantly physical disabilities). The models were optimized by observing the lowest root mean squared error in the combination of feature selections within the MS training cohort.

### 2.5 GBM model validation

With the final optimized models, two validations methods were performed: (1) 10-fold cross-validation and (2) an independent cohort validation. 10-fold cross-validation reuses the training cohort data by randomly partitioning the data into an “internal” training set (90% of the total training cohort) and validation set (10% of the total training cohort) on different iterations. The model then tests prediction accuracy of the withheld samples. However, the independent cohort validation strategy utilizes a completely new dataset that was removed before model development. This allows true evaluation of the models’ performances in predicting clinical scores.

Correlations between measured and predicted clinical scores were evaluated using linear regression models in R Studio version 4.1.0 (Team, 2015; Team, 2021), reporting R^2, Spearman Rho (the relationship strength of the predicted and measured scores) from R CRAN Package: stats (Team, 1969) and rstatix (Kassambara, 2021); and Concordance Correlation

Coefficient (CCC; the degree of reliability in the method when comparing two measurements of the same variable) from R CRAN Package: DescTools (Signorell et al., 2021).

Additional materials and methods are included as Supplemental text.

## 3. RESULTS

### 3.1 Regressing out physiological confounders

We regressed out effects of physiological confounders based on internal HV cohort as described in Methods and exemplified in Figure 2. For ventricular CSF volume, the multiple linear regression model selected only effects of age^2 and intracranial volume from the 6 tested confounders. For some other CNS structures, the multiple linear regression models were more complex. But consistent with published studies, we saw no independent effects of BMI on any CNS volume.

### 3.2 MS-specific residual effects of age and gender on CNS volumes

Several remaining confounding effects on GM volumes were noted in MS cohort. All GM structures demonstrated same paradoxical trend of increasing GM volumes with age of MS patients. This relationship reached statistical significance (i.e., Bonferroni adjusted p<0.004) for caudate (R^2=0.04, p=0.002; Supplementary Figure S2A), cerebrum GM (R^2=0.06, p<0.001, Supplementary Figure S2B), putamen (R^2=0.08, p<0.001; Supplementary Figure S2C) and supratentorial GM (R^2=0.06, p<0.001, Supplementary Figure S2D). Since these residual effects were not seen in non-MS cohort (Supplementary Figure S2), they are MS-specific and are driven by non-physiologically low GM volumes in young patients. Although the unadjusted volumes of these GM structures decrease with age (consistent with other longitudinal studies), the paradoxical positive residual effect of age in MS emerges after subtracting physiological confounders because the slope of age-related decrease is higher in HV and non-MS cohorts.

In contrast to GM where younger MS patients exhibited more profound non-physiological atrophy compared to older MS patients, we observed residual linear loss of supratentorial WM (R^2=0.10; p<0.001; Supplementary Figure S3B) and normal appearing WM (R^2=0.09; p<0.001; Supplementary Figure S3A) with MS aging, paralleled by significant residual increase in ventricular (but not sulcal) CSF volume (R^2=0.04; p=0.002, Supplementary Figure S4B). This progressive non-physiological WM loss was MS specific (i.e., it was not observed in non-MS cohort) and affected also the cerebellum WM even though the residual correlation of cerebellum WM with age in MS became non-significant after Bonferroni adjustment. In fact, the large non-MS cohort (n=160) behaved identically to HV regarding all WM volumes, while it exhibited non-physiological atrophy in some GM structures (including thalamus, Supplementary Figure S4A) and non-physiological increase in ventricular volume (Supplementary Figure S4B).

We conclude that subtracting effects of physiological confounders on volumes of CNS structures identified non-physiological loss of GM volumes in young MS patients and MS specific, progressive non-physiological loss of WM volumes.

### 3.3 Differences in unadjusted and confounder-adjusted MRI features between HV and MS

As shown in Supplementary Figure S1, HV cohort was significantly younger than MS (i.e., Median 40.4 years vs Median=52.4 years; p=1.24e-7). Because age exerted significant effects on many CNS volumes, subtracting the variance explained by age diminished differences between HV and MS cohorts in most CNS volumetric biomarkers (Figure 3). The greatest decrease in effect size was seen for thalamus, caudate and brain parenchymal fraction (BPFr) - consistent with the reported strong effect of physiological aging on these CNS structures.

Unexpectedly, we observed increase in effect sizes of WM volumes on differentiating MS from HV after confounder adjustment: Supratentorial WM and normal appearing WM (which are highly correlated – see Figure 4) but marginally also cerebellum WM.

**Figure 3.**
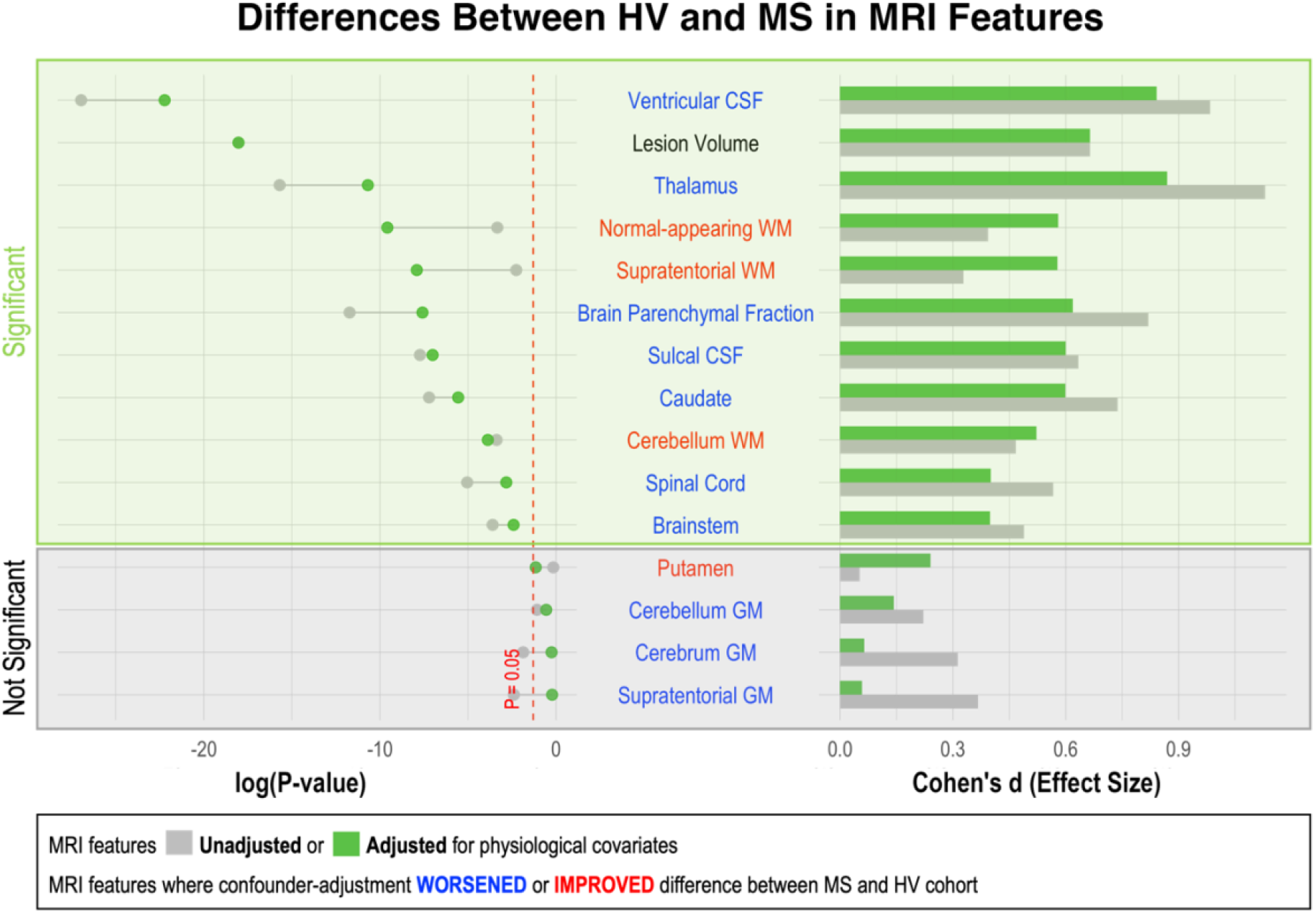
Effect of adjustment for physiological confounders on measured differences in MRI volumetric features between HV and MS patients. Right side of the figure shows effect sizes of unadjusted (gray bars) and confounder-adjusted (green bars) volumetric MRI features to differentiate MS from HV. Effect sizes are shown as Standardized Main Difference (i.e., Cohen’s d). Left side of the figure shows the effect of applied confounder adjustment with log p-value. MRI features where confounder adjustment decreased the difference between MS and HV are highlighted in blue, whereas those MRI features where confounder adjustment increased the difference are highlighted in red. 11 out of 15 MRI features differentiated MS from HV with statistical significance. The difference between MS and HV was partially driven by confounder differences in majority of these MRI features (6/10). Thus, the applied adjustment decreased the ability of these features to differentiate MS from HV. On the other hand, three measurements of white matter volume (supratentorial WM, normal-appearing WM and cerebellum WM) enhanced their ability to differentiate MS from HV after confounder adjustment.

**Figure 4.**
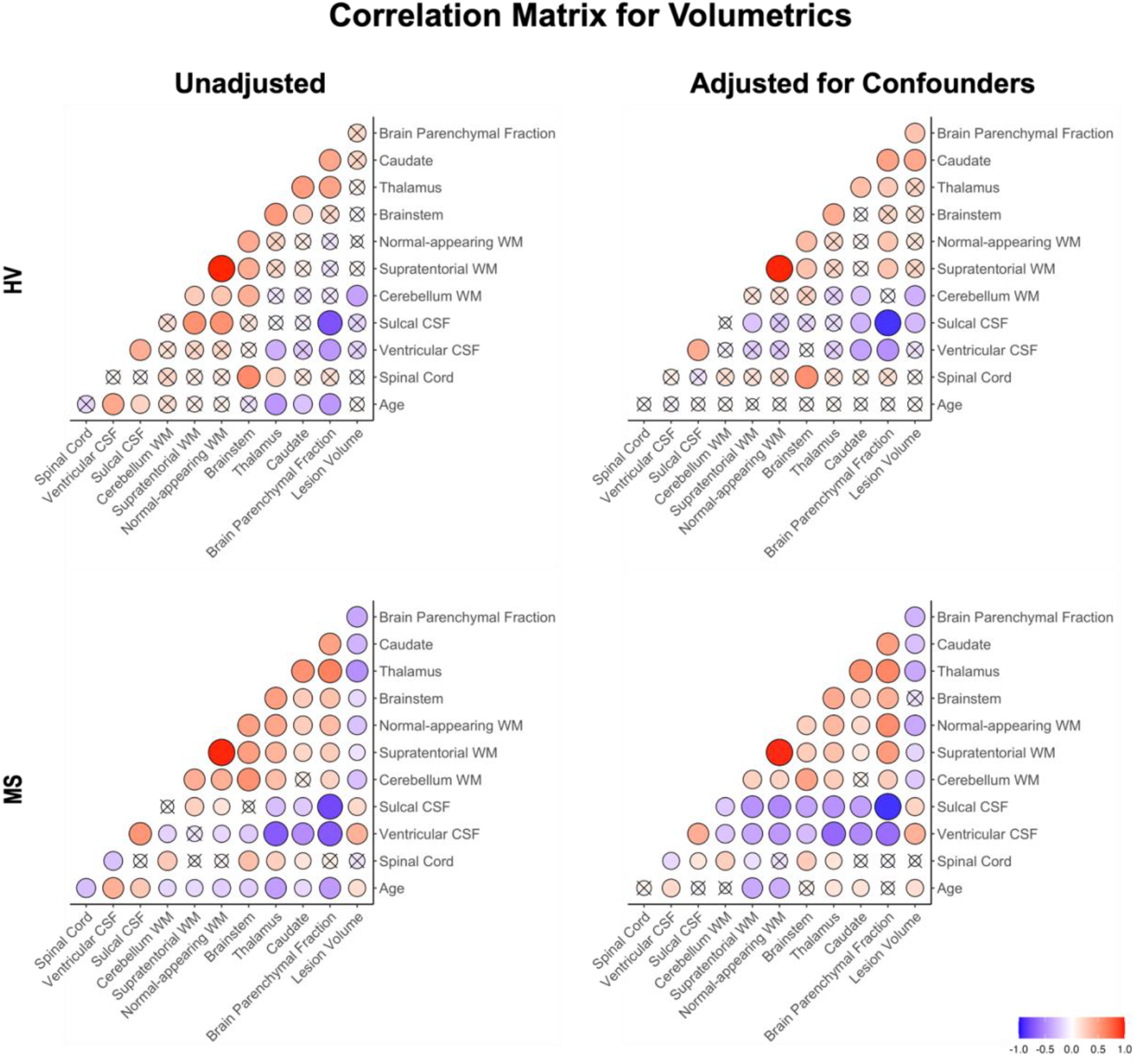
Correlation matrix of unadjusted and confounder-adjusted MRI features. The top row shows correlations in HV cohort. Bottom row shows correlations in MS training cohort. Left panel shows correlations of unadjusted MRI features. Right panel shows correlations of confounder-adjusted MRI features. Correlations that are not statistically significant are marked as (x), positive correlations are marked in red and negative in blue colors. The size of the circle corresponds to Spearman’s correlation coefficient. Confounder adjustment eliminated correlations of MRI features with age, while it generally strengthened correlations of brain parenchymal fraction (BPFr) with remaining MRI features.

Confounder adjustment affected correlations between MRI features (Figure 4). As expected, it eliminated correlations of MRI biomarkers with age in the HV cohort. But the adjustment also enhanced correlations of BPFr with most brain volumes in both HV and MS cohorts. Finally, confounder adjustments strengthened correlations of brain WM volumes and ventricular CSF with remaining MRI features in MS.

### 3.4 Univariate correlations of MRI features with clinical outcomes

Figure 5 and Supplementary Table S1 show univariate Spearman correlation coefficients and p-values of unadjusted and HV confounder-adjusted MRI volumes with clinical outcomes in the MS training and validation cohorts. We observed that for physical disability outcomes (EDSS, CombiWISE and NeurEx), age exerted comparable or higher effect size on clinical outcomes than the strongest MRI biomarkers. This was not true for SDMT, where brain lesion volume and ventricular CSF had strongest correlations.

**Figure 5.**
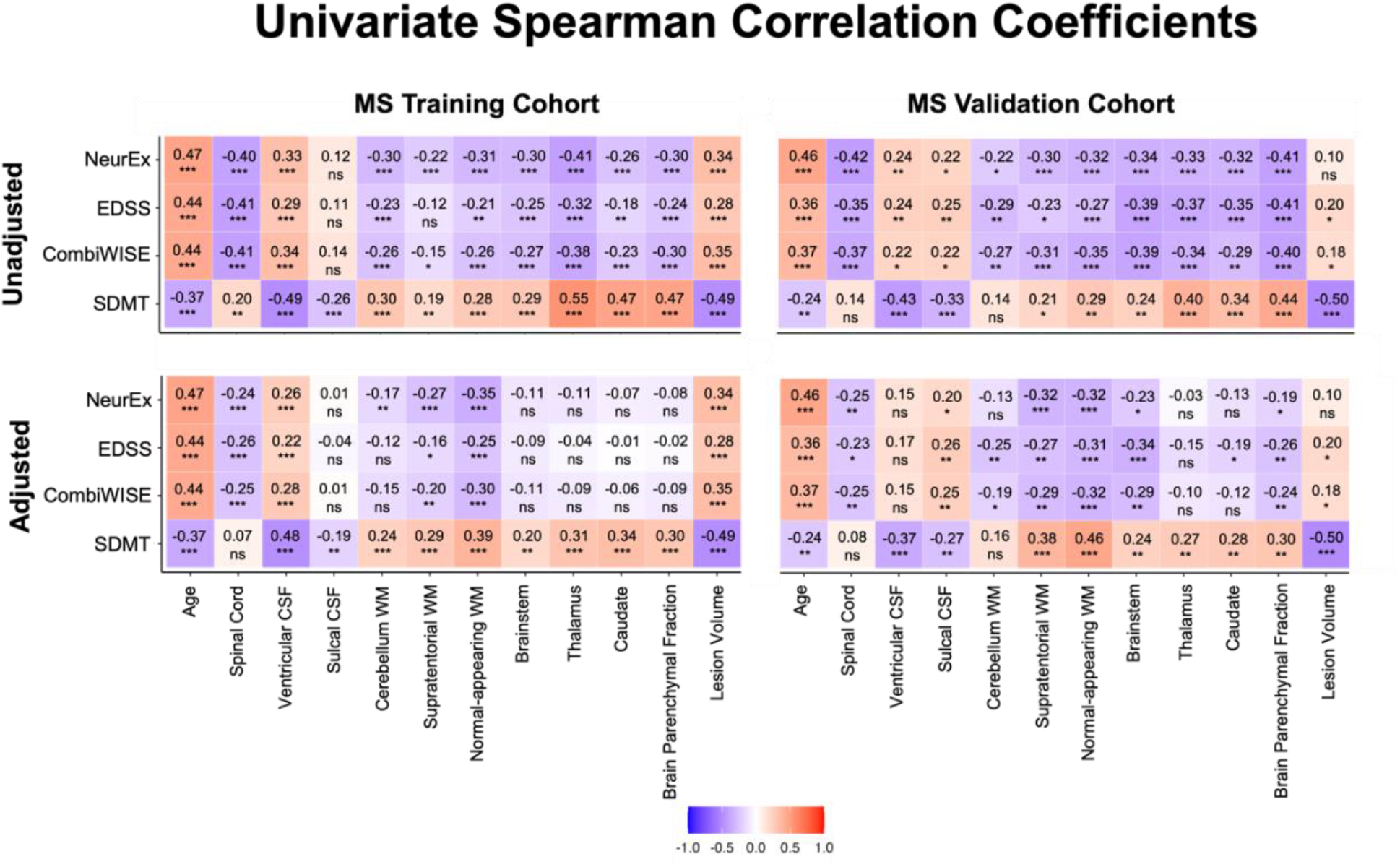
Unilateral correlation matrix of unadjusted and confounder-adjusted MRI features with disability outcomes. The top row shows unilateral correlations of disability outcomes with unadjusted MRI features. Left panel shows correlations of MS training cohort and right panel shows correlations of MS validation cohort. Bottom row shows analogous correlations with confounder-adjusted MRI features. Correlations that are not statistically significant are marked as (ns). Positive correlations are marked in red and negative in blue colors. Correlation p-values indicate statistical significance (*P < 0.05, **P < 0.01, and ***P < 0.001). Additional descriptive statistics are shown in Supplementary Table S1.

As would be expected from strong univariate correlations of age with disability outcomes and with MRI volumes, subtracting age effects significantly diminished correlations of CSF volumes, GM volumes and even BPFr with clinical outcomes.

However, confounder adjustment strengthened correlation of WM volumes with all clinical outcomes (in both cohorts). In fact, WM volumes became the highest correlated MRI biomarkers with physical disability outcomes (NeurEx and CombiWISE) and NAWM became the second highest (after lesion volume) correlating biomarker with SDMT in the validation cohort after confounder adjustment.

We conclude that adjustment for physiological confounders reproducibly strengthened correlations of supratentorial WM volumes with MS clinical outcomes.

### 3.5 Models from MRI volumetric data adjusted for physiological confounders achieve stronger effect sizes in predicting MS clinical outcomes in the independent validation cohort

Unadjusted and confounder-adjusted MRI features that achieved statistical significance in differentiating HV and MS cohorts were inputted into ML-based models of four clinical outcomes. Because age and gender exerted significant influence on clinical outcomes and their effects were subtracted from confounder-adjusted MRI volumes, we added these demographic variables into the model that used adjusted MRI volumes. The full statistical results from all models are in Supplementary Table S2.

For the three outcomes of physical disability (i.e., CombiWISE, EDSS and NeurEx), we observed stronger effect sizes for unadjusted models (Figure 6A and Supplementary Table S2). But for SDMT, the confounder-adjusted model slightly outperformed the unadjusted model.

**Figure 6.**
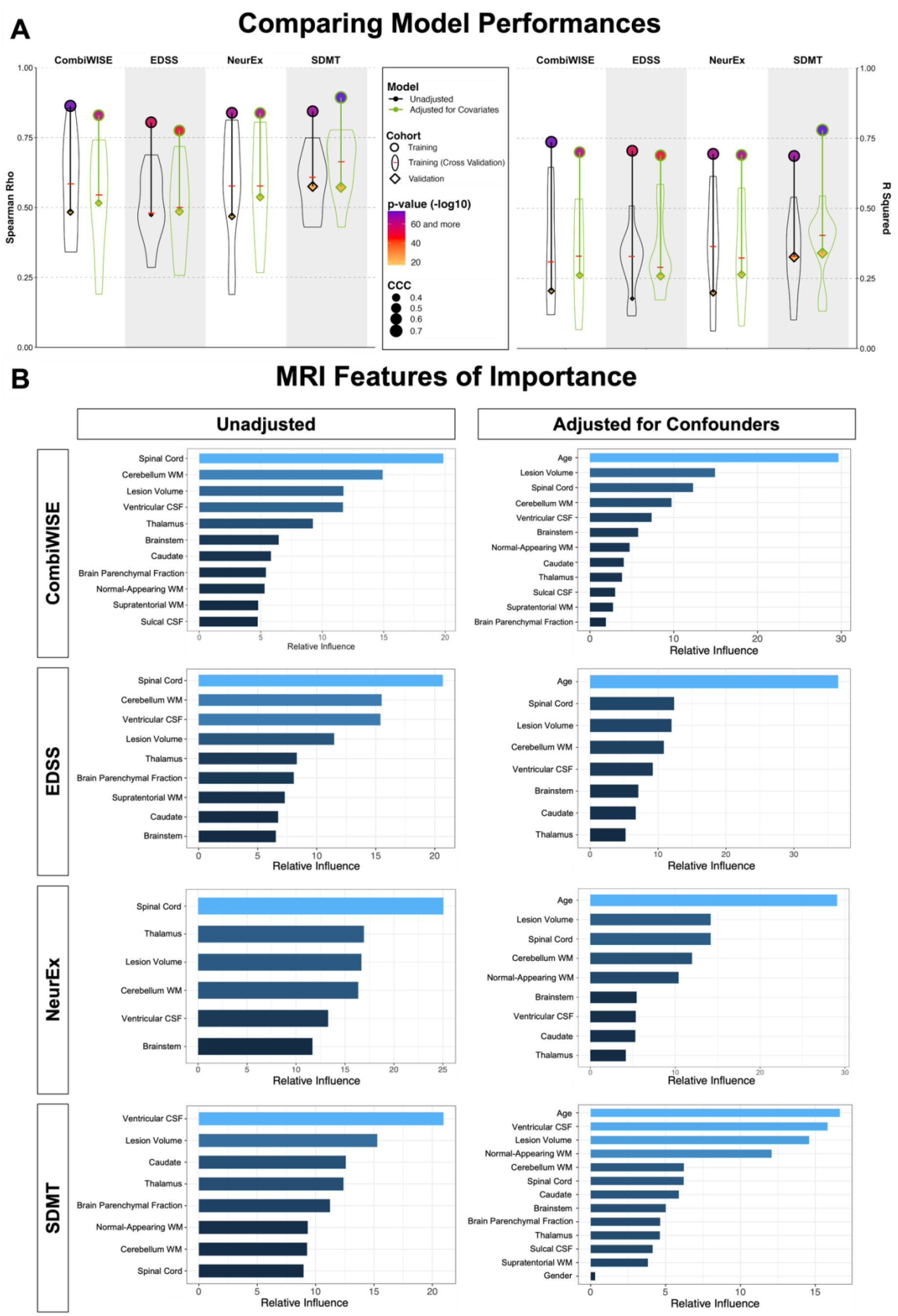
Summary of model performances. **A**. The left panel shows Spearman Rho correlation coefficients while right panel shows coefficient of determination (R^2) for the same 8 models. For each outcome (arranged in vertical columns), the unadjusted models are on the left side outlined in black, and the confounder-adjusted models are on the right side, outlined in green. The model performance in the training cohort is shown as circle; the distribution of cross-validation (i.e., re-using of the training cohort) results is shown as a violin plot with the median marked as a red horizontal line; the independent validation cohort performance is shown as diamond. The size of the symbols (i.e., circle and diamond) correspond to the Linh’s concordance coefficient (CCC). Finally, the color of each symbol represents p-value (-log10) with lower p-values displayed in purple color and higher p-values in orange in accordance with the heatmap displayed in the figure legend. Unequivocal and robust decrease in model’s performance is seen from training cohort to cross-validation to independent validation. Detailed statistics of the model performances are shown in Supplementary Table S2. **B**. For each clinical outcome modelled (presented in rows) we show number of features selected in the final model(s) arranged in descending order of variable importance. The unadjusted models are displayed on the left, while confounder-adjusted models are displayed on the right.

This hierarchy between models was generally preserved in the training cohort 10-fold cross-validation results, shown in Figure 6A as violin plots with medians marked by red cross-sectional line. Cross-validation have broad distribution of effect sizes in comparison to the full training cohort: e.g., while training cohort Spearman correlation coefficients (Rho) were higher than 0.75 for all eight models, cross-validation results ranged from Rho 0.2 to Rho 0.8, showcasing the poor estimate of model’s performance depending on the training cohort splits. Nevertheless, cross-validation medians had uniformly lower effect sizes than the training cohort; the decrease in effect sizes was substantial (between 40-60% for R^2).

The independent validation cohort achieved effect sizes consistently below the cross-validation medians, showcasing that the training cohort, including training cohort cross-validation, significantly over-estimate models’ performance. Nevertheless, all eight models validated with very low p-values (all below 9E-8; Supplementary Table S2).

The most surprising observation was that confounder-adjusted models consistently outperformed models from unadjusted features in the independent validation cohort. The absolute difference in R^2 values between unadjusted and adjusted models was up to R^2=0.08 (i.e., EDSS unadjusted model achieved R^2=0.18, while adjusted model had R^2=0.26; Supplementary Table S2). This represents relative improvement of 30%.

We conclude that training cohort results are poor predictors of the model’s performance in the independent validation cohort: they consistently and dramatically over-estimate final effect sizes. The stronger validation performance of confounder-adjusted models supports our hypothesis that effects of aging and other physiological covariates on MRI volumes represents noise when predicting which MS-related CNS tissue damage contributes to clinical disability.

This does not indicate that age does not play role in MS. In fact, age was selected as the strongest feature in all confounder-adjusted models (Figure 6B), implying that age is the most important determinant of MS-related disability. Our results simply support the hypothesis that age exerts effects on CNS structures by (mostly) different mechanisms in physiological aging and in MS. By separating the effect of age on MS from its effect on brain structures during natural aging, we built more reliable models that predict MS-associated disability with higher effect sizes in the independent validation cohort.

## 4. DISCUSSION

The goal of this project was to gain understanding of MS-driven volumetric MRI changes and to examine if computational models of confounder-adjusted MRI volumes reproducibly predict different clinical outcomes.

Before we discuss our findings, we want to point out following limitations: our HV cohort is relatively small and the Lesion-TOADS automated segmentation algorithm, implemented from the inception of our natural history protocol, has relatively narrow adoption. Despite the differences in cohort sizes and the segmentation algorithms, our observations are aligned with published literature, both for HV (Potvin et al., 2016) and MS (Ontaneda et al., 2021; Simpson-Yap et al., 2021). Specifically, although we tested six confounders, only age (and age^2), gender and total intracranial volume influenced volumetric brain MRI features with effect sizes comparable to those reported by others (Potvin et al., 2016). Second, we did not examine the interaction between confounders because others (Potvin et al., 2016) found that these had negligible effects. To facilitate broader use and potential external validation of our results, we collaborated with QMENTA to implement publicly available Lesion-TOADS algorithm on their platform. The other important limitation is that by the virtue of being national referral center, our patient population may be skewed towards subject with more aggressive disease. However, this limitation applies to all MS imaging studies, as these are performed almost uniformly in tertiary academic centers. Finally, our comparably high effect sizes in predicting MS disability could be, at least partially due to lower technical variability by virtue of a single center study that uses standardized scanning protocols. Indeed, MAGNIMS investigators reported that inter-scanner/protocol confounders may explain >10% of variance (Cole et al., 2020).

Notwithstanding these limitations, our study achieved its aims. Our GBM models from confounder-adjusted MRI features predicted four MS disability scales with high statistical significance and good effect sizes in the independent validation cohort. These models slightly outperformed analogous models derived from unadjusted MRI features and significantly outperformed any single MRI biomarker.

Considering that approximately 50% of variance in brain volumes can be explained by confounders, subtracting these confounders significantly diminished the difference between MS and HV cohorts (which differed in age) and decreased univariate correlations with MS clinical outcomes (which positively correlate with age). Ignoring such large effects of confounders on MRI volumes overestimates how well the measured change reflects MS progression.

Unexpectedly, confounder adjustment increased effect sizes (and lowered p-values) for WM MRI volumes: for both supratentorial WM and normal appearing WM (which correlate strongly with each other), but also for cerebellum WM, where the improvement was marginal. The confounder-adjusted WM volumes had comparable effect size to BPFr in differentiating MS from HV.

We conclude that after subtracting the effects of natural aging and sexual dimorphism, WM pathology (represented by formation of MS lesions associated with atrophy of deep GM structures and enlargement of ventricles) is the dominant effect of MS visible on conventional brain MRI. The resulting correlation matrix of confounder-adjusted MRI volumes supports this conclusion by demonstrating relatedness of MS-induced changes in all aforementioned structures. Similarly, we saw that these brain structures were selected by most GBM models, with predictably higher influence of WM volumes in the models based on confounder-adjusted biomarkers.

Although brain structures with higher effect sizes in differentiating MS from HV were more important in GBM models of disability, this was not true for SC CSA. C1-2 SC CSA had marginal effect size and p-value in differentiating MS from HV, but it had relatively high univariate correlations with disability outcomes and, concordantly, was selected by all GBM models of physical disability (i.e., EDSS, CombiWISE, and NeurEx) as leading MRI feature. These results are consistent with clinical observations that MS involvement of SC is the dominant driver of physical disability, especially ambulation. We believe that relatively weak effect size in differentiating MS from HV for SC CSA resides in the technical difficulty of scanning such small structure. We did not have dedicated SC MRIs for most subjects, and artifacts due to CSF pulsation, heart beats and breathing are likely accentuated by using brain coil, rather than dedicated SC coil with protocol optimized for SC imaging.

Additionally, age was selected as the strongest variable in confounder-adjusted models. This poses a question whether we gained anything by adjusting MRI features for confounders. First, confounder adjustment did enhance validation performance of the models. Second, in unilateral correlations, age correlates with disability outcomes with effect sizes comparable to strongest MRI biomarkers. However, this does not mean that effect of age on MS disability is pathophysiologically identical to normal aging. For example, age effects on immunity may become relevant only in patients with intrathecal inflammation. Separating age from its confounding effect on MRI biomarkers may lead to models that are more responsive to MS treatment effects, a potential hypothesis for future longitudinal studies.

Thanks to a recent meta-analysis of 302 papers describing models of MS clinical outcomes (Liu et al., 2022), we can compare our results with other published MRI biomarker-based models. Using an associated website that allow users to dynamically explore this rich dataset, we identified 40 papers that used MRI biomarkers to model EDSS as ordinal scale and reported p-values, and 20 papers that reported effect sizes as R^2. Many studies overestimate effect sizes from using small cohorts or failing to implement methodological design that limit bias (Button et al., 2013; Ioannidis, 2005, 2008). Thus, the meta-analysis scored methodological rigor of reviewed studies by grading 7 criteria: 1. Blinding, 2. Defined strategy to deal with outliers, 3. Explanation of missingness, 4. Adjustment for confounders, 5. Number of comparisons made and whether p-values were adjusted, 6. Presence of controls and 7. Validation (cross-validation of the training cohort versus independent validation cohort). The median numbers of criteria fulfilled by published studies that modelled EDSS was 2, and the majority studied less than 100 MS patients. Only 38% of studies adjusted p-values for multiple comparisons, and studies that did not adjust performed up to 500 comparisons. The studies with the highest methodological rigor and largest cohorts originated from the MAGNIMS consortium. MAGNIMS study of effects of GM brain volumes on differentiating MS (n=961) from HV (n=203) and on disability prediction found negative association between deep GM (β =-0.71; p<0.0001) and cortical GM (β =-0.22; p<0.0001) and EDSS. Unfortunately, R^2 was not reported. For much smaller studies that reported R^2, the range was 0.7 to 0.05 in the training cohort. Only one study reported cross-validation/OOB results (Cordani et al., 2021) and achieved R^2=0.19 (p-value range from <0.001 to 0.04) in RRMS (n=250) and R^2=0.16 (p-value range from 0.02 to 0.04) in PMS (n=114). Current study fulfills 7/7 criteria of methodological rigor and is the only study that includes independent validation cohort. For the confounder-adjusted EDSS model, the training cohort R^2 is 0.69 (p=3.8e-43). Median cross-validation R^2 is 0.29 and the independent validation cohort R^2 is 0.26 (p=2.4e-08). To our knowledge, this is the strongest reported effect size for predicting EDSS as ordinal scale from quantitative MRI biomarkers in the literature thus far.

Analogously, we identified 12 studies that used MRI predictors for modeling SDMT. 11/12 reported p-values and 5/12 reported R^2. The median methodological rigor score was 2/7 and most cohorts were smaller than 100 subjects (the largest had 151 subjects). Only 16.7% adjusted for multiple comparisons and the studies that did not adjust p-values performed up to 100 comparisons. No studies reported cross-validation or independent validation. The reported R^2 (for the training cohorts) ranged from 0.62-0.3. Again, our confounder-adjusted SDMT model achieved R^2=0.78 (p=7.3e-75) with independent validation R^2=0.34 (p=2.9e-11), which represent the best performance among published studies.

We have observed that cross-validation generates broad range of results that encompass the effect sizes of the independent validation cohort, but the median of cross-validation results is still overly optimistic. This is consistent with our extensive observations using independent validation in all our projects (Boukhvalova et al., 2019; Liu et al., 2022; Masvekar et al., 2021; Messan et al., 2022; Pham et al., 2021). Therefore, while cross-validation should be included in all modeling studies, the independent validation must be considered a gold standard. This is extremely important, as only 15% of published MS models used any validation strategy and only 8% used independent validation.

In conclusion, our final models outperform correlations of any single MRI biomarker with clinical outcomes. These models are therefore likely more sensitive imaging outcomes and should be explored in future clinical trials, especially when targeting older subjects with progressive MS who no longer form acute MS lesions. The MAGNIMS consortium has available datasets to quantify possible loss of sensitivity due to inter-scanner/inter-protocol variance on such ML-based models in comparison to a single MRI biomarker.

## Supporting information

All Supplementary Figures

## Data Availability

All data produced in the present study are available upon reasonable request to the authors.

## Data Availability

All raw data and script used for all analyses will be provided after being accepted.

## Author Contribution

YK and MV performed MRI scan processing, volumetric analyses, and maintained the cloud based QMENTA platform to generate brain volumetric data. YK performed all analyses detailed in the paper, generated the figures, and contributed to writing the paper. PK exported all clinical scores used in correlation analyses and supervised in the development of GBM models. BB construed the project conceptually, guided and supervised all aspects of the study, and contributed to the writing of the paper and figure creations. All authors critically reviewed and edited the manuscript.

## Acknowledgements

We are grateful for all study participants as well as everyone in the Clinical Immunology and Microbiology (LCIM) for their support and feedbacks.

