## Supplementary material for "Confounder-adjusted MRI-based predictors of multiple sclerosis disability": All Supplementary Figures

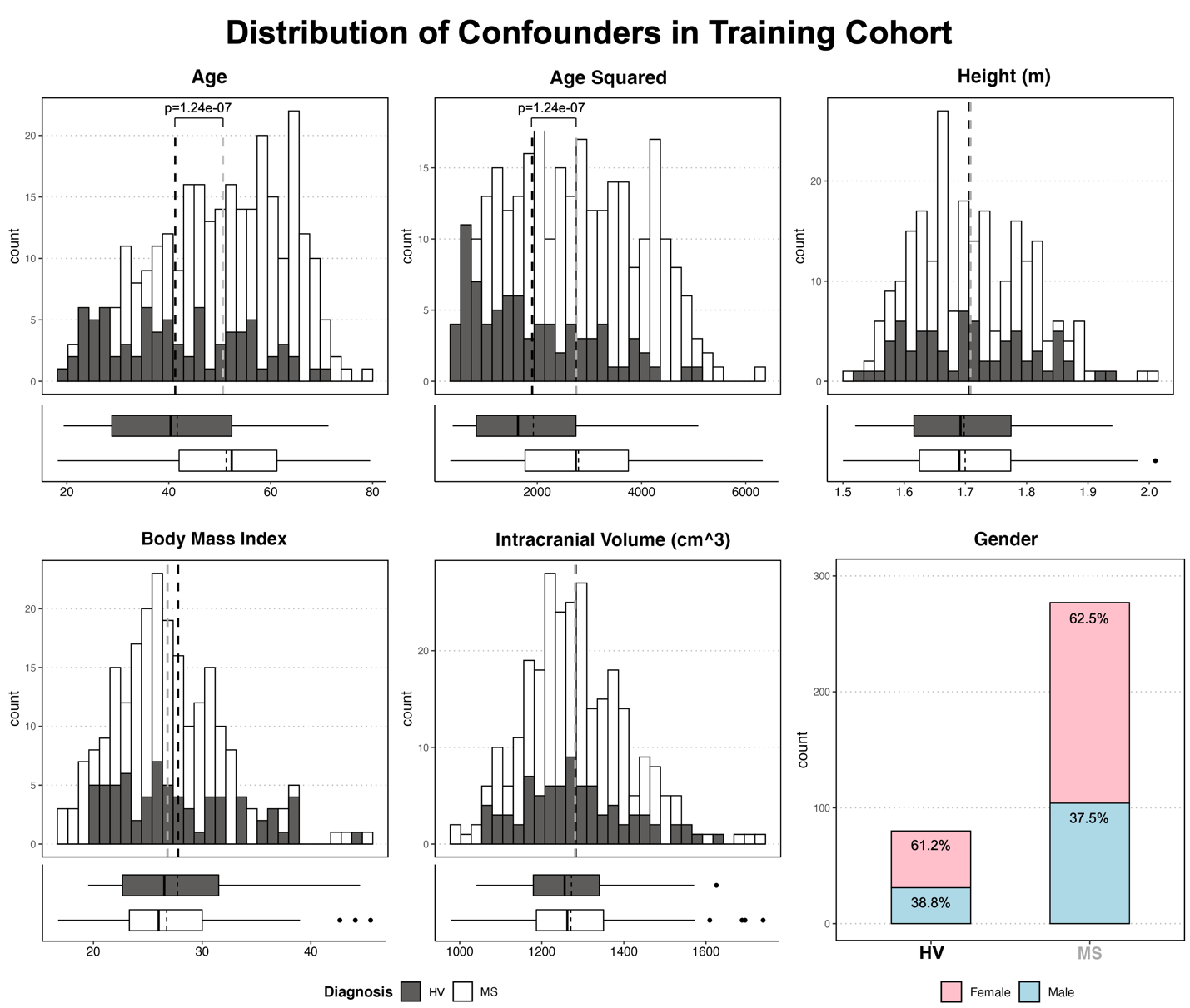

**Supplementary Figure S1.** **Differences in cohort characteristics between healthy volunteers (HV) and MS patients.** Distribution of five continuous confounders is shown as histograms with HV cohort in black and MS cohort in while colors. The proportions and absolute numbers of males and females in HV and MS cohorts are displayed as a bar chart with females show in pink and males in blue. There are no significant differences between HV and MS cohort in all confounders except age (and age^2).

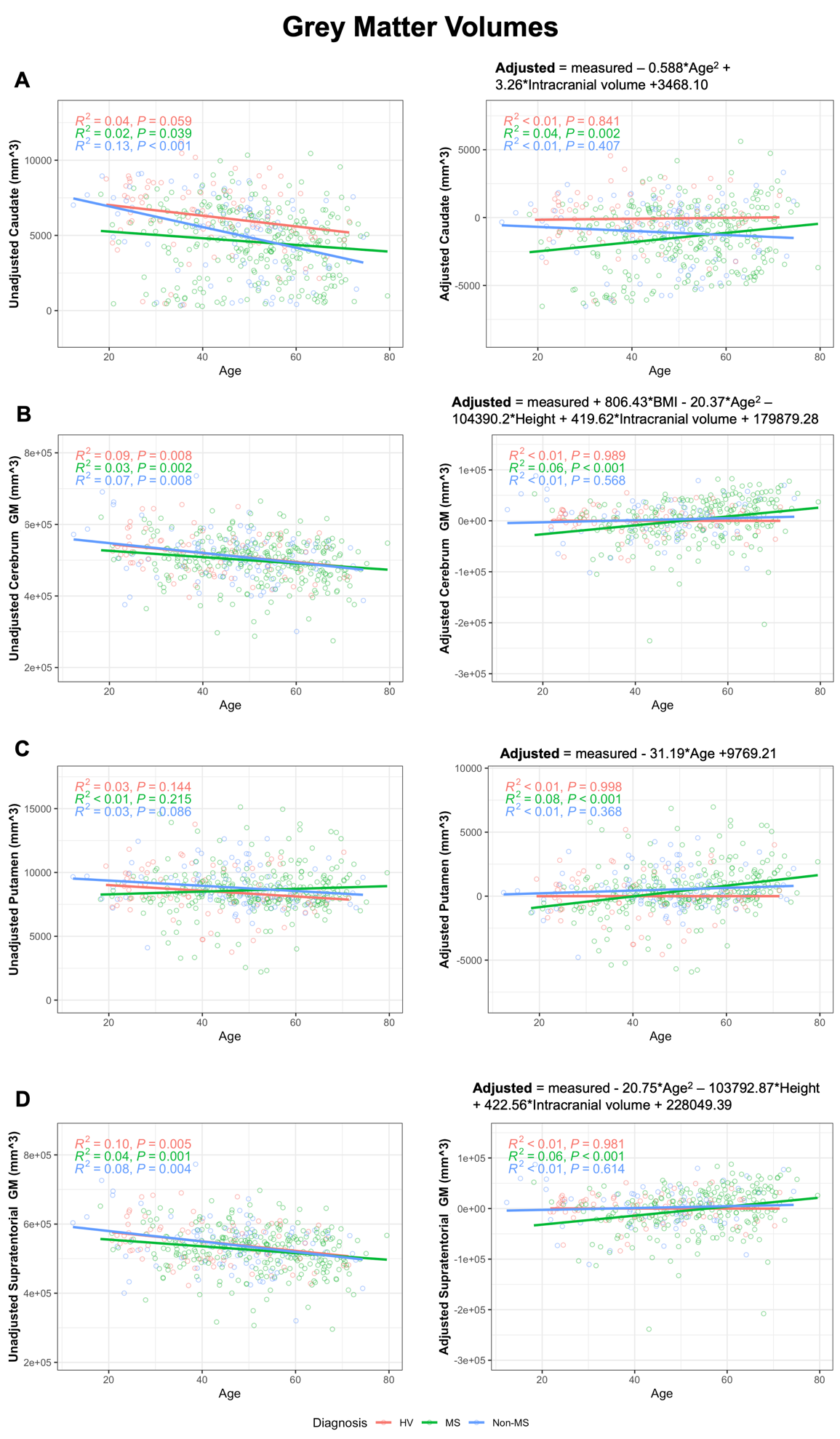

**Supplementary Figure S2. Grey matter volumes with residual variance with age within training cohort.** Left panels all show unadjusted grey matter volumes while right panels show confounder-adjusted grey matter volumes. Red line and points indicate HV cohort while blue line and points indicate MS cohort. Adjustment equations respective to MRI feature are shown on top of the plot showing adjusted MRI volumes.

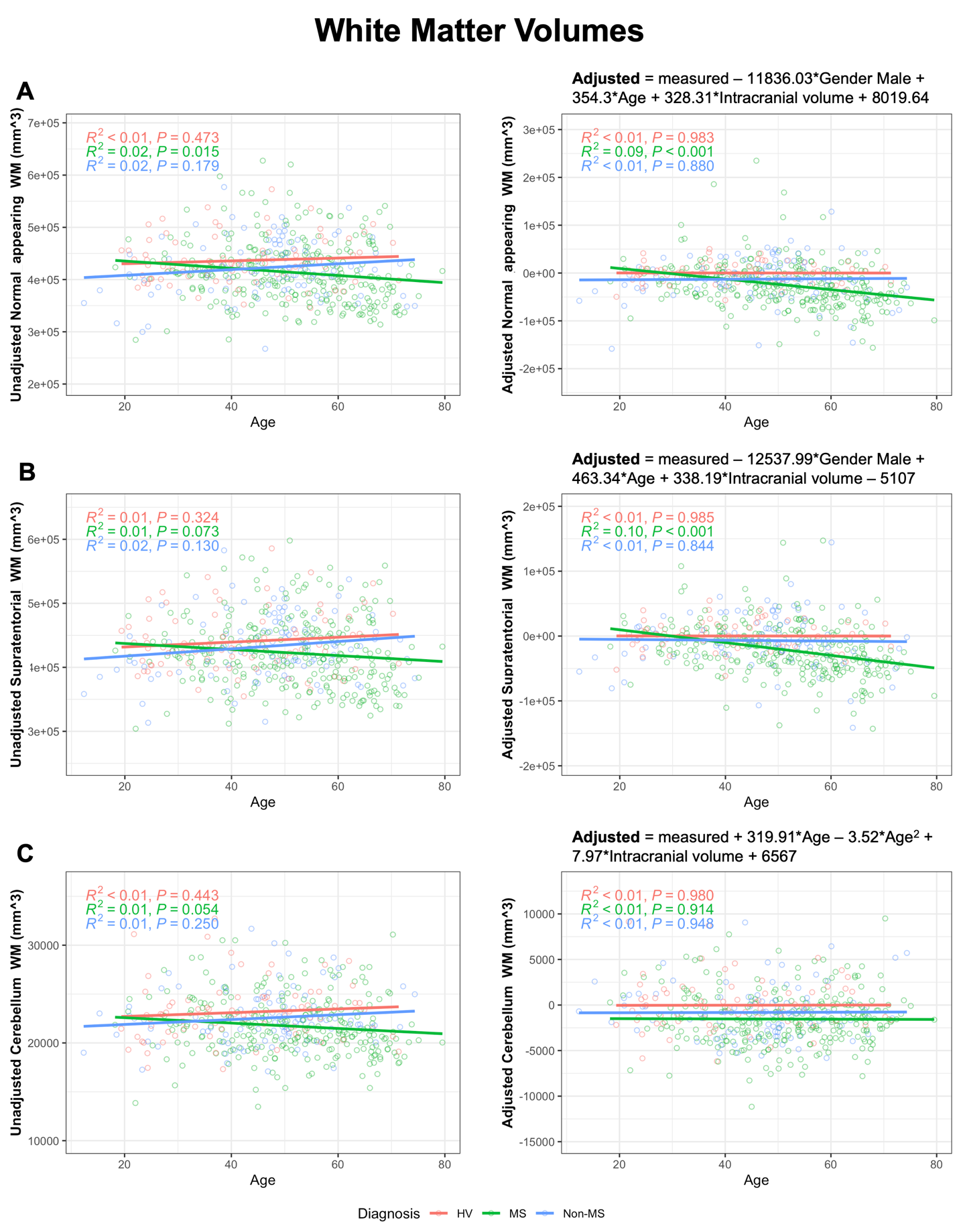

**Supplementary Figure S3.** Left panels all show unadjusted white matter volumes while right panels show confounder-adjusted white matter volumes. Red line and points indicate HV cohort while blue line and points indicate MS cohort. Adjustment equations respective to MRI feature are shown on top of the plot showing adjusted MRI volumes.

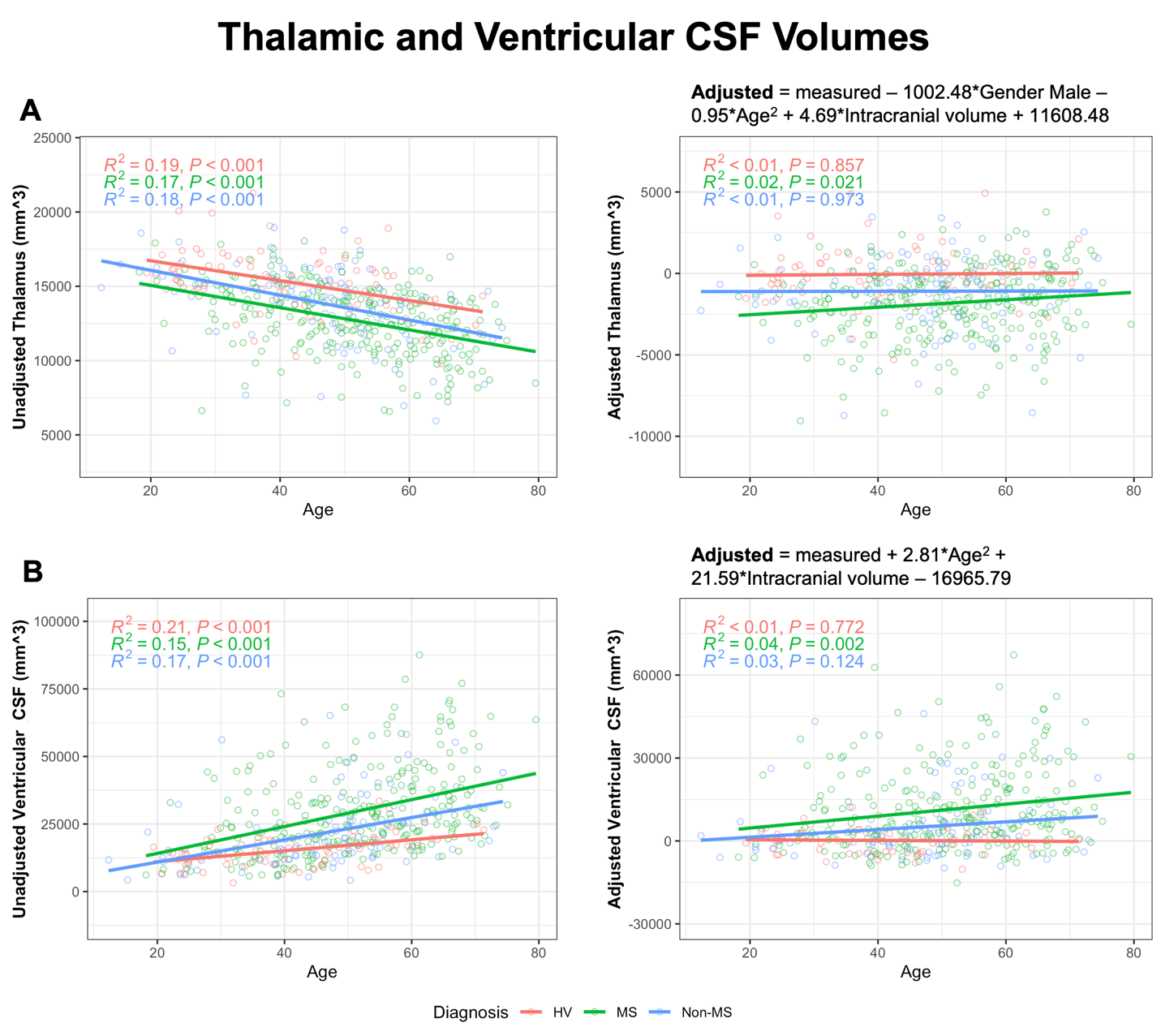

**Supplementary Figure S4.** Left panels all show unadjusted thalamic and ventricular CSF volumes while right panels show confounder-adjusted volumes. Red line and points indicate HV cohort while blue line and points indicate MS cohort. Adjustment equations respective to MRI feature are shown on top of the plot showing adjusted MRI volumes.

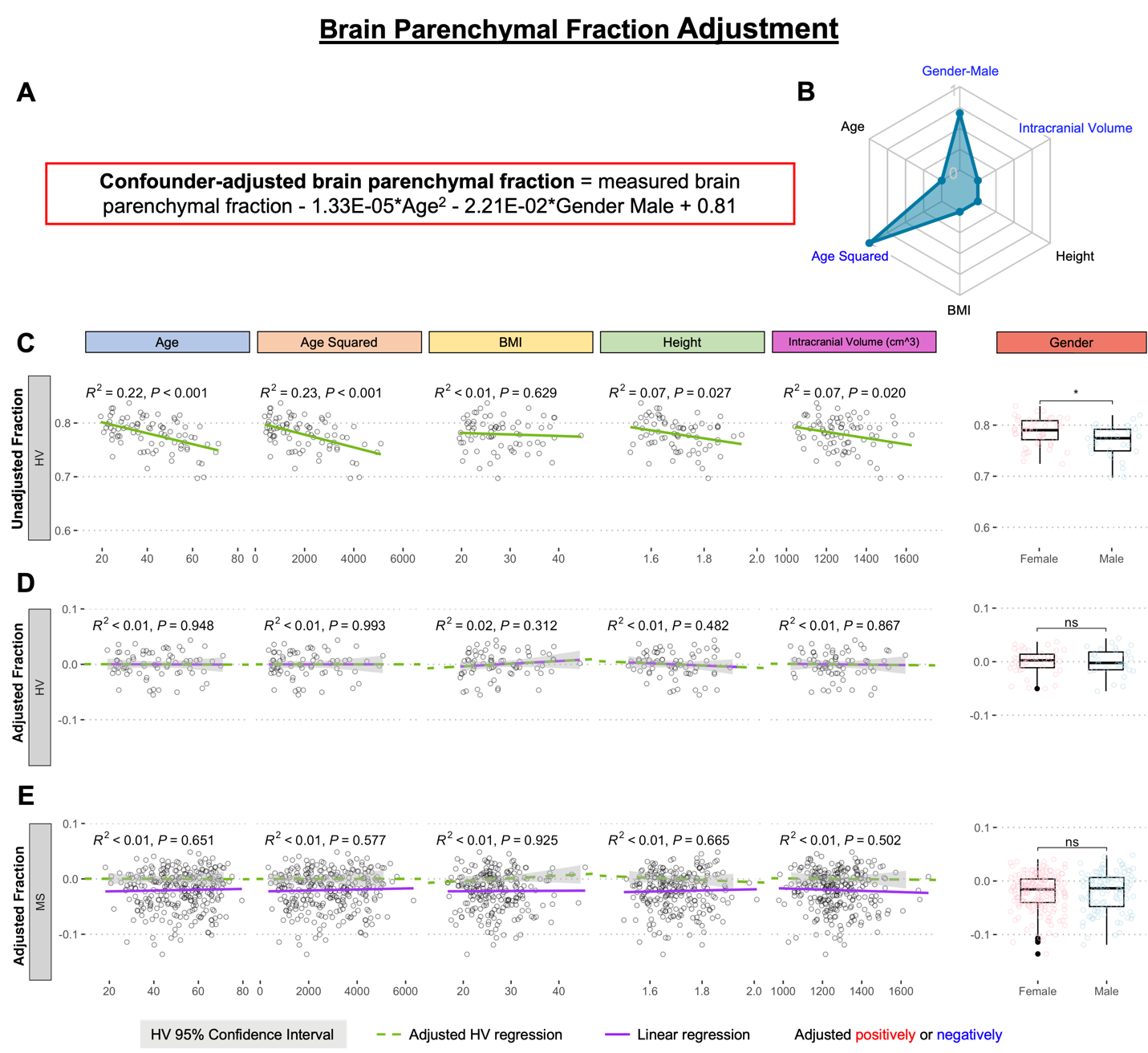

**Supplementary Figure S5. Adjustment of single MRI biomarker (i.e., brain parenchymal fraction) for six physiological confounders. A.** The final equation for the confounder-adjusted brain parenchymal fraction is shown at the top in red outline. This equation was derived from multiple linear regression models as described in the Method section. **B.** In the top right corner is resulting radar chart that show proportional weights of the applied confounder adjustment, with confounders with lowest weights (i.e., the innermost circle) representing zeros. **C.** Univariate linear regression models between each tested confounder on x-axis (first 5 graphs) or gender (sixth graph) and measured brain parenchymal fraction on the y-axis in 80 healthy volunteers (HV). **D.** Same univariate regressions in the HV cohort after applying adjustment formula show no remaining effect of confounders. **E.** Applying HV-derived adjustment formula to MS cohort shows no residual effect.

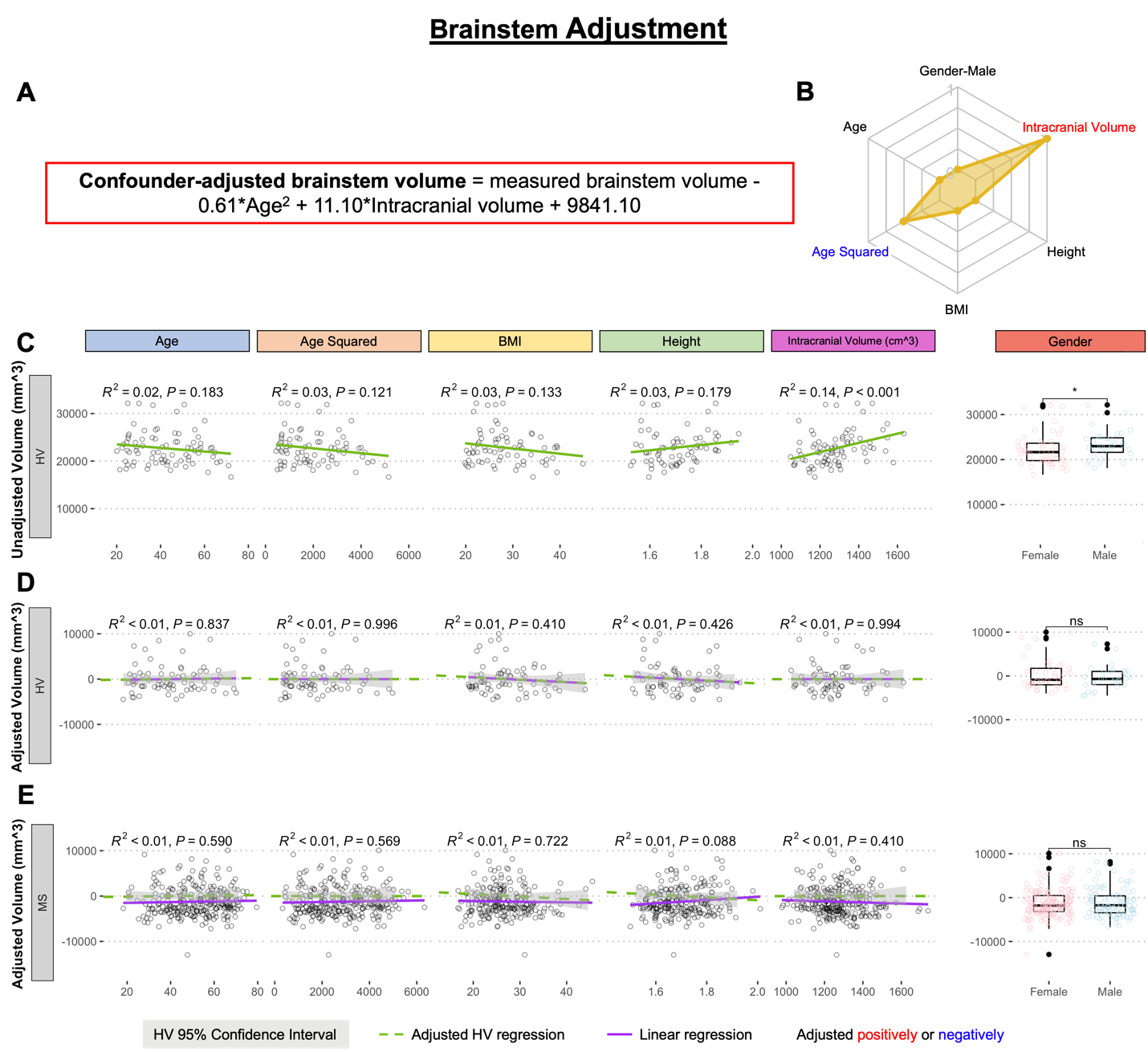

**Supplementary Figure S6. Adjustment of single MRI biomarker (i.e., brainstem volume) for six physiological confounders. A.** The final equation for the confounder-adjusted brainstem volume is shown at the top in red outline. This equation was derived from multiple linear regression models as described in the Method section. **B.** In the top right corner is resulting radar chart that show proportional weights of the applied confounder adjustment, with confounders with lowest weights (i.e., the innermost circle) representing zeros. **C.** Univariate linear regression models between each tested confounder on x-axis (first 5 graphs) or gender (sixth graph) and measured brainstem volume on the y-axis in 80 healthy volunteers (HV). **D.** Same univariate regressions in the HV cohort after applying adjustment formula show no remaining effect of confounders. **E.** Applying HV-derived adjustment formula to MS cohort resulted in no residual effect.

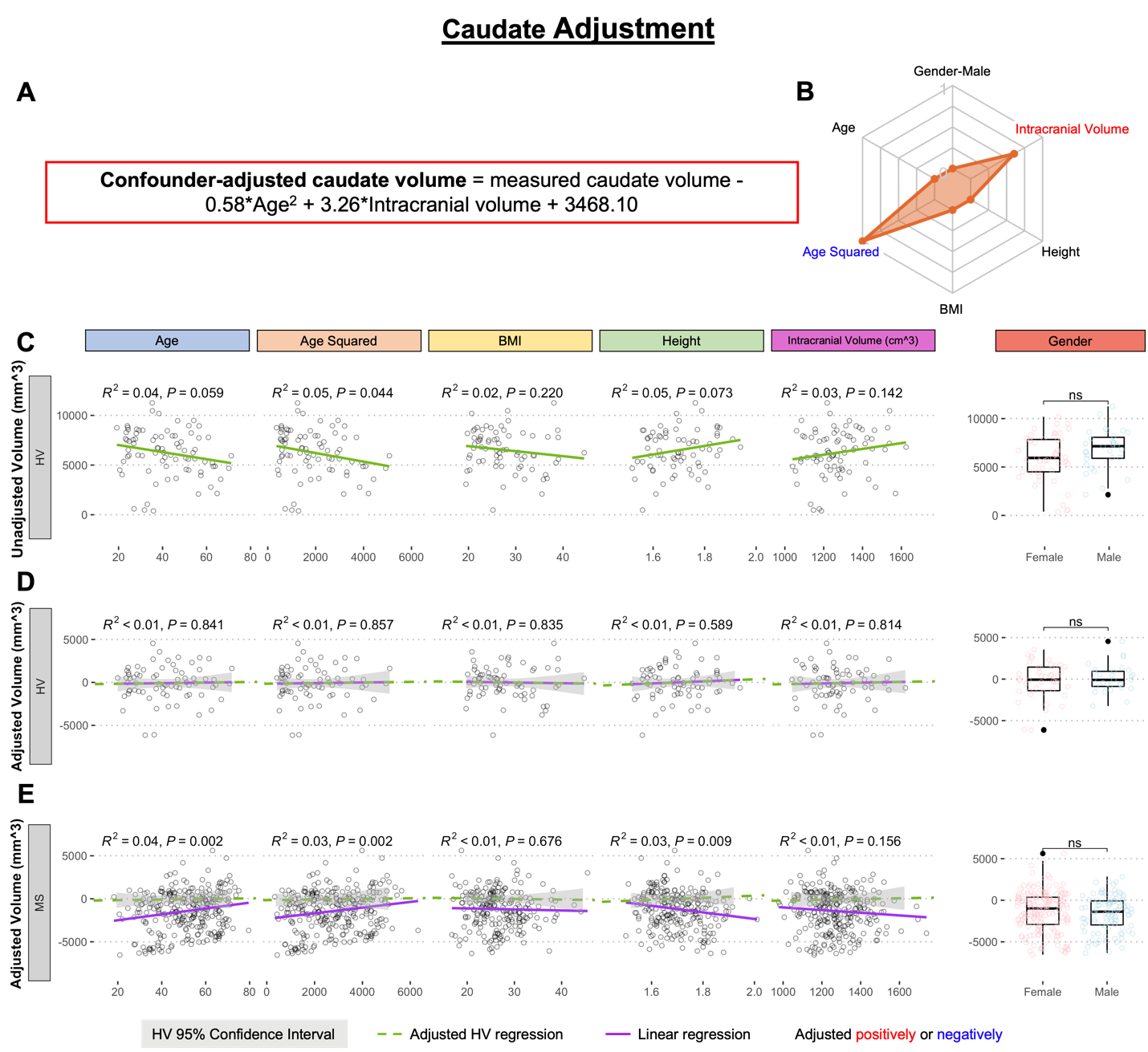

**Supplementary Figure S7. Adjustment of single MRI biomarker (i.e., caudate volume) for six physiological confounders. A.** The final equation for the confounder-adjusted caudate volume is shown at the top in red outline. This equation was derived from multiple linear regression models as described in the Method section. **B.** In the top right corner is resulting radar chart that show proportional weights of the applied confounder adjustment, with confounders with lowest weights (i.e., the innermost circle) representing zeros. **C.** Univariate linear regression models between each tested confounder on x-axis (first 5 graphs) or gender (sixth graph) and measured caudate volume on the y-axis in 80 healthy volunteers (HV). **D.** Same univariate regressions in the HV cohort after applying adjustment formula show no remaining effect of confounders. **E.** Applying HV-derived adjustment formula to MS cohort shows residual effect of age and age^2, when considering Bonferroni adjustment for multiple comparisons (i.e., p<0.05/12 = p<0.004).

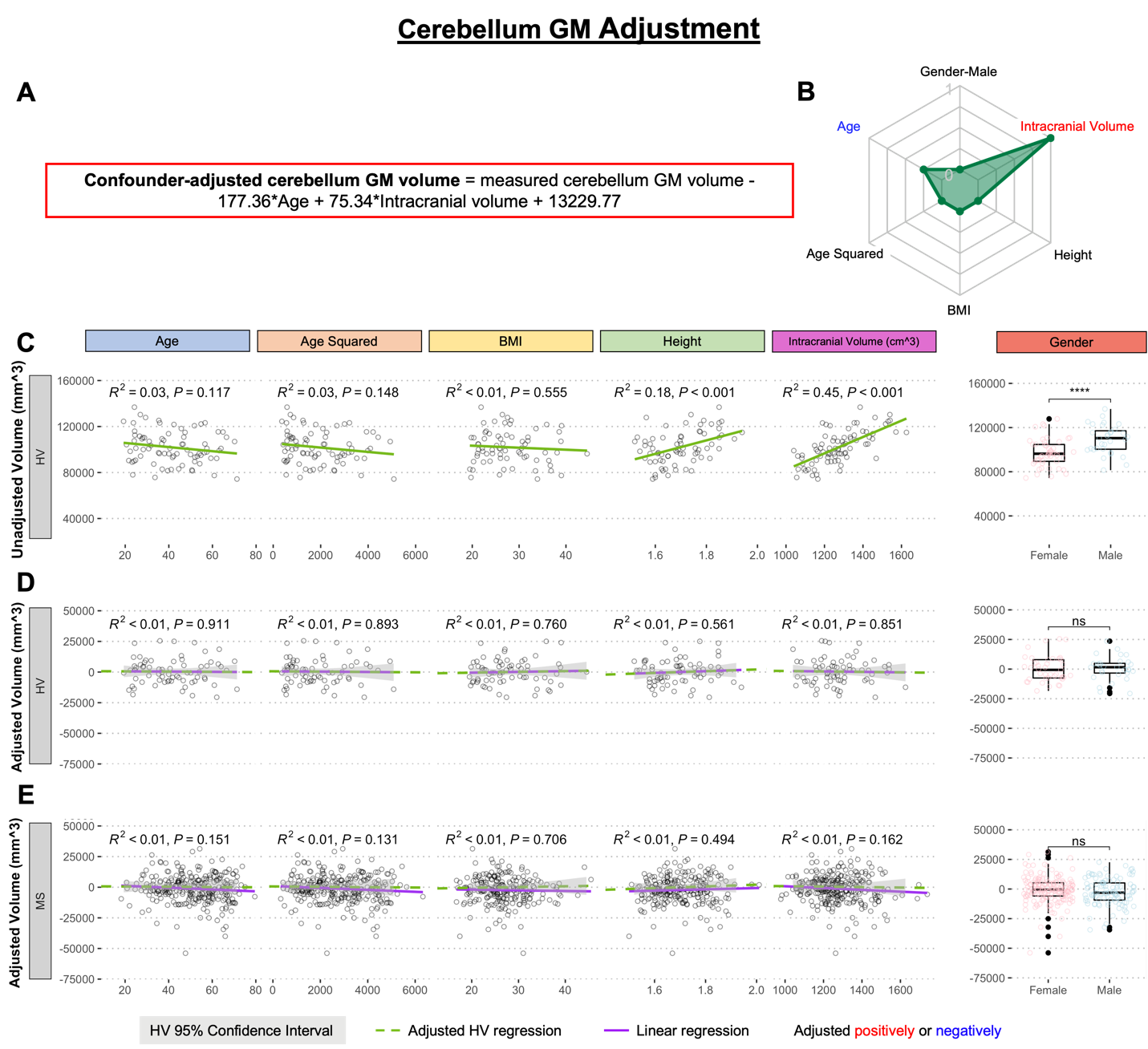

**Supplementary Figure S8. Adjustment of single MRI biomarker (i.e., cerebellum GM volume) for six physiological confounders. A.** The final equation for the confounder-adjusted cerebellum GM volume is shown at the top in red outline. This equation was derived from multiple linear regression models as described in the Method section. **B.** In the top right corner is resulting radar chart that show proportional weights of the applied confounder adjustment, with confounders with lowest weights (i.e., the innermost circle) representing zeros. **C.** Univariate linear regression models between each tested confounder on x-axis (first 5 graphs) or gender (sixth graph) and measured cerebellum GM volume on the y-axis in 80 healthy volunteers (HV). **D.** Same univariate regressions in the HV cohort after applying adjustment formula show no remaining effect of confounders. **E.** Applying HV-derived adjustment formula to MS cohort resulted in no residual effect.

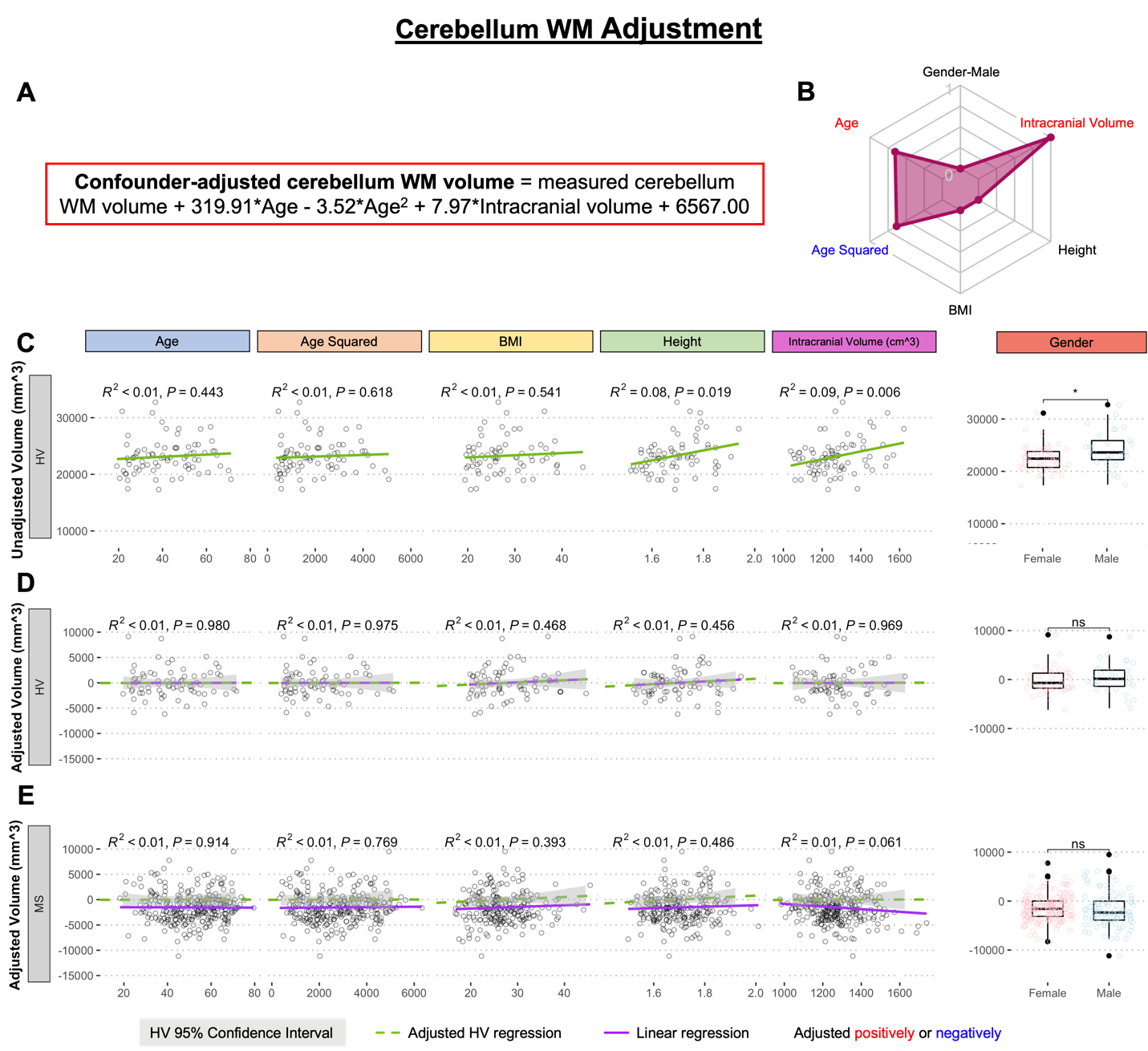

**Supplementary Figure S9. Adjustment of single MRI biomarker (i.e., cerebellum WM volume) for six physiological confounders. A.** The final equation for the confounder-adjusted cerebellum WM volume is shown at the top in red outline. This equation was derived from multiple linear regression models as described in the Method section. **B.** In the top right corner is resulting radar chart that show proportional weights of the applied confounder adjustment, with confounders with lowest weights (i.e., the innermost circle) representing zeros. **C.** Univariate linear regression models between each tested confounder on x-axis (first 5 graphs) or gender (sixth graph) and measured cerebellum WM volume on the y-axis in 80 healthy volunteers (HV). **D.** Same univariate regressions in the HV cohort after applying adjustment formula show no remaining effect of confounders. **E.** Applying HV-derived adjustment formula to MS cohort resulted in no residual effect.

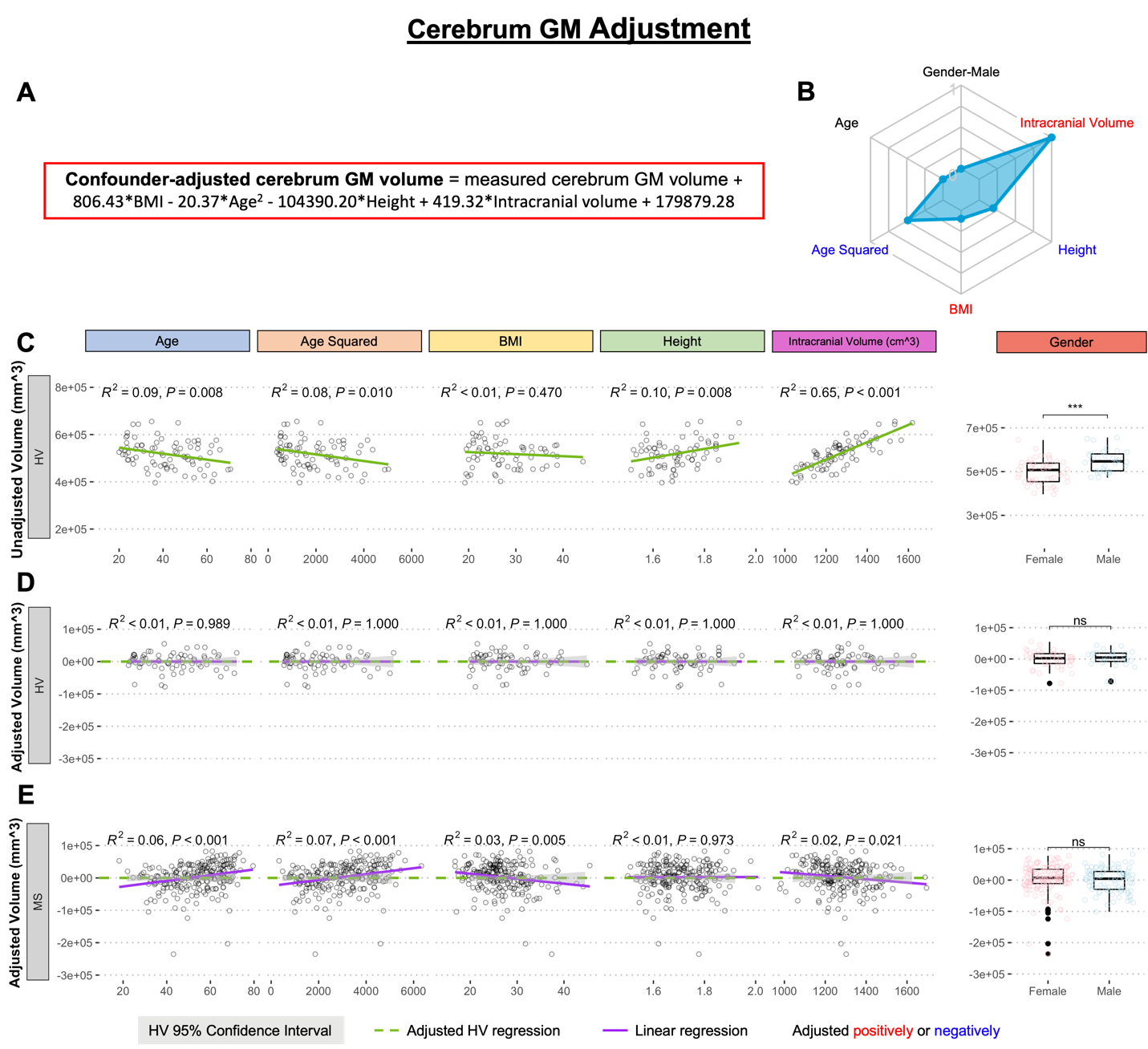

**Supplementary Figure S10. Adjustment of single MRI biomarker (i.e., cerebrum GM volume) for six physiological confounders. A.** The final equation for the confounder-adjusted cerebrum GM volume is shown at the top in red outline. This equation was derived from multiple linear regression models as described in the Method section. **B.** In the top right corner is resulting radar chart that show proportional weights of the applied confounder adjustment, with confounders with lowest weights (i.e., the innermost circle) representing zeros. **C.** Univariate linear regression models between each tested confounder on x-axis (first 5 graphs) or gender (sixth graph) and measured cerebrum GM volume on the y-axis in 80 healthy volunteers (HV). **D.** Same univariate regressions in the HV cohort after applying adjustment formula show no remaining effect of confounders. **E.** Applying HV-derived adjustment formula to MS cohort shows residual effect of age and age^2 when considering Bonferroni adjustment for multiple comparisons (i.e., p<0.05/12 = p<0.004).

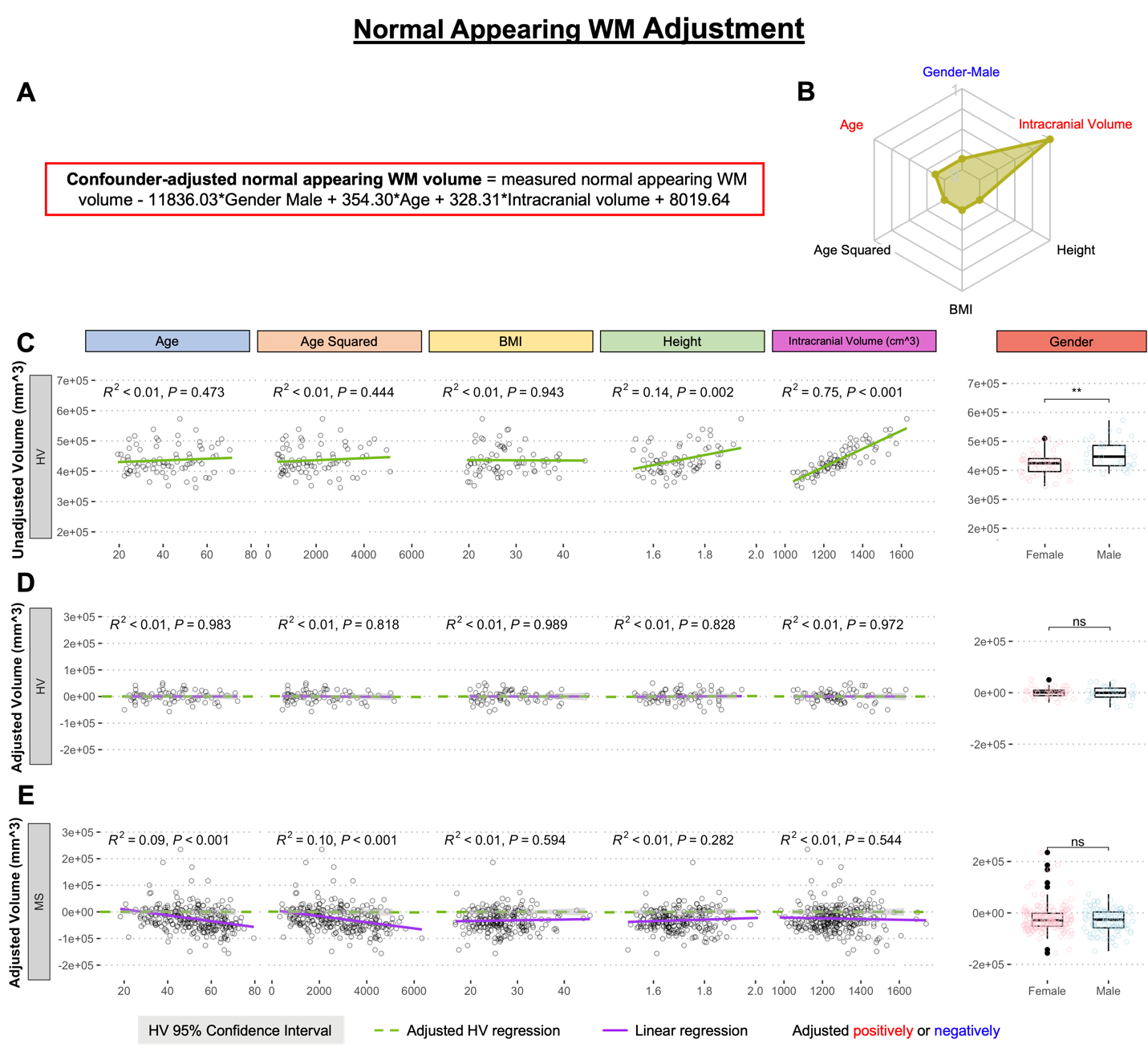

**Supplementary Figure S11. Adjustment of single MRI biomarker (i.e., normal-appearing WM volume) for six physiological confounders. A.** The final equation for the confounder-adjusted normal-appearing WM volume is shown at the top in red outline. This equation was derived from multiple linear regression models as described in the Method section. **B.** In the top right corner is resulting radar chart that show proportional weights of the applied confounder adjustment, with confounders with lowest weights (i.e., the innermost circle) representing zeros. **C.** Univariate linear regression models between each tested confounder on x-axis (first 5 graphs) or gender (sixth graph) and measured normal-appearing WM volume on the y-axis in 80 healthy volunteers (HV). **D.** Same univariate regressions in the HV cohort after applying adjustment formula show no remaining effect of confounders. **E.** Applying HV-derived adjustment formula to MS cohort shows residual effect of age and age^2.

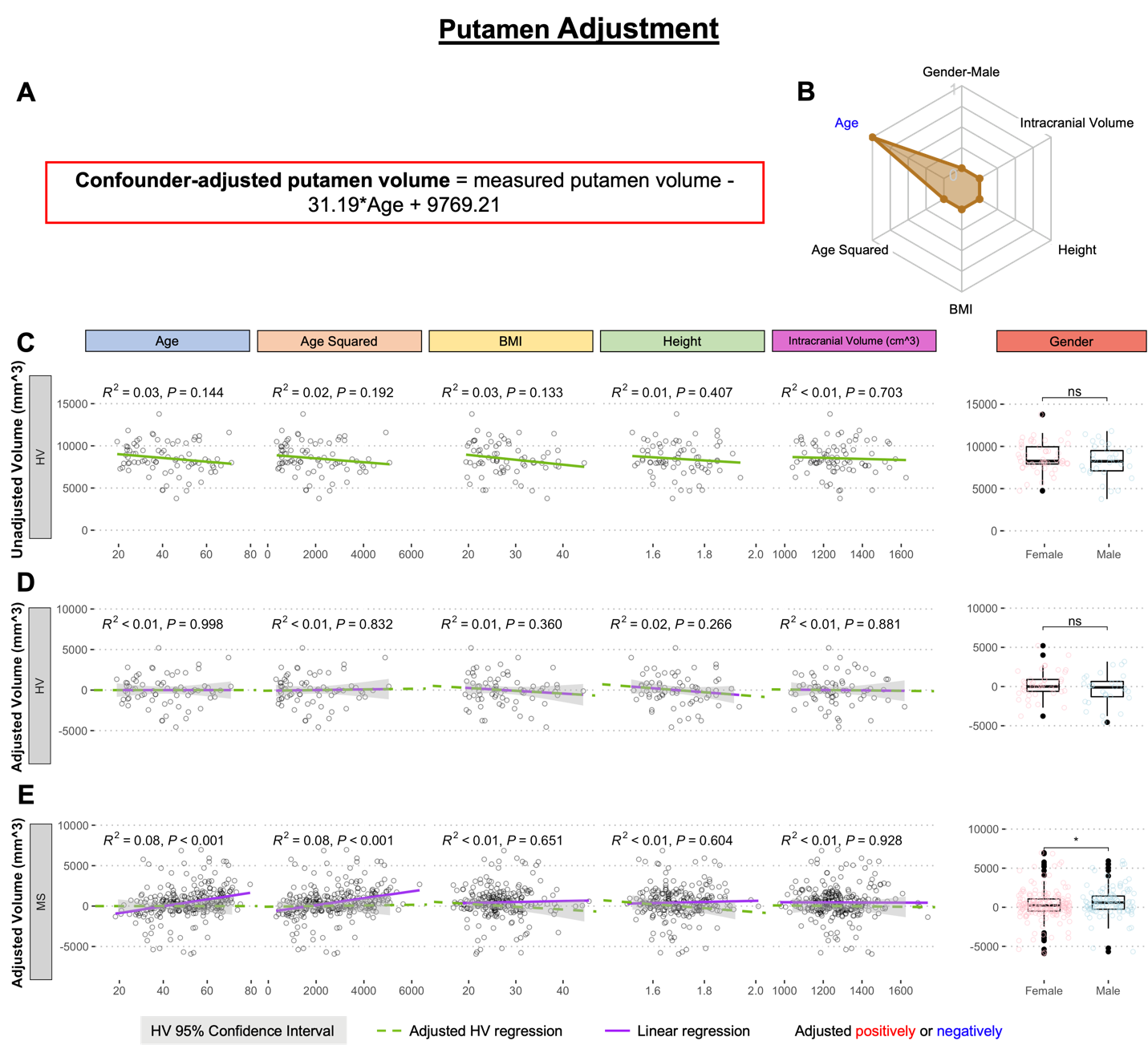

**Supplementary Figure S12. Adjustment of single MRI biomarker (i.e., putamen volume) for six physiological confounders. A.** The final equation for the confounder-adjusted putamen volume is shown at the top in red outline. This equation was derived from multiple linear regression models as described in the Method section. **B.** In the top right corner is resulting radar chart that show proportional weights of the applied confounder adjustment, with confounders with lowest weights (i.e., the innermost circle) representing zeros. **C.** Univariate linear regression models between each tested confounder on x-axis (first 5 graphs) or gender (sixth graph) and measured putamen volume on the y-axis in 80 healthy volunteers (HV). **D.** Same univariate regressions in the HV cohort after applying adjustment formula show no remaining effect of confounders. **E.** Applying HV-derived adjustment formula to MS cohort shows residual effect of age and age^2, when considering Bonferroni adjustment for multiple comparisons (i.e., p<0.05/12 = p<0.004).

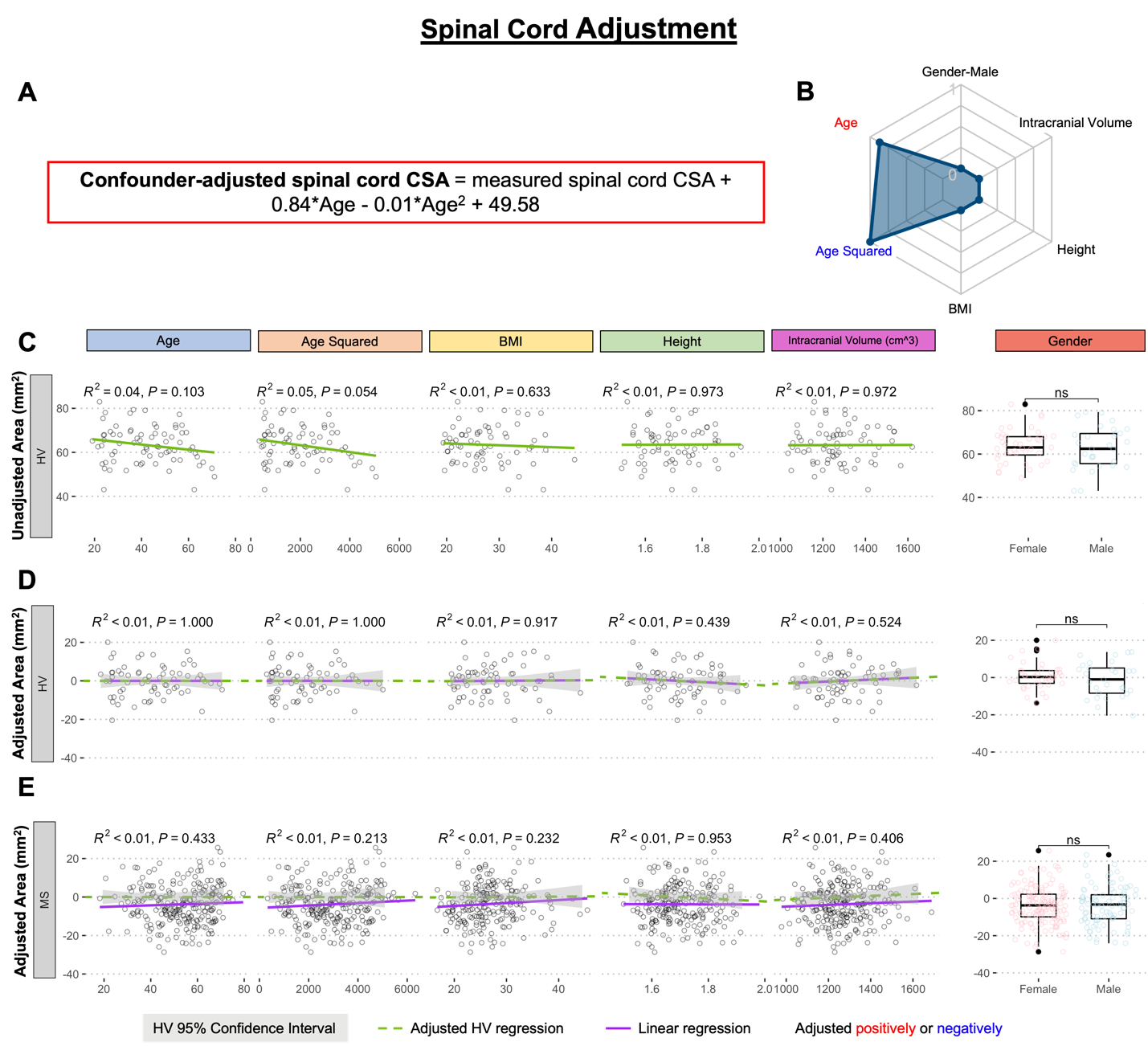

**Supplementary Figure S13. Adjustment of single MRI biomarker (i.e., spinal cord average cross section area) for six physiological confounders. A.** The final equation for the confounder-adjusted spinal cord cross section area (CSA) is shown at the top in red outline. This equation was derived from multiple linear regression models as described in the Method section. **B.** In the top right corner is resulting radar chart that show proportional weights of the applied confounder adjustment, with confounders with lowest weights (i.e., the innermost circle) representing zeros. **C.** Univariate linear regression models between each tested confounder on x-axis (first 5 graphs) or gender (sixth graph) and measured spinal cord CSA on the y-axis in 80 healthy volunteers (HV). **D.** Same univariate regressions in the HV cohort after applying adjustment formula show no remaining effect of confounders. **E.** Applying HV-derived adjustment formula to MS cohort resulted in no residual effect.

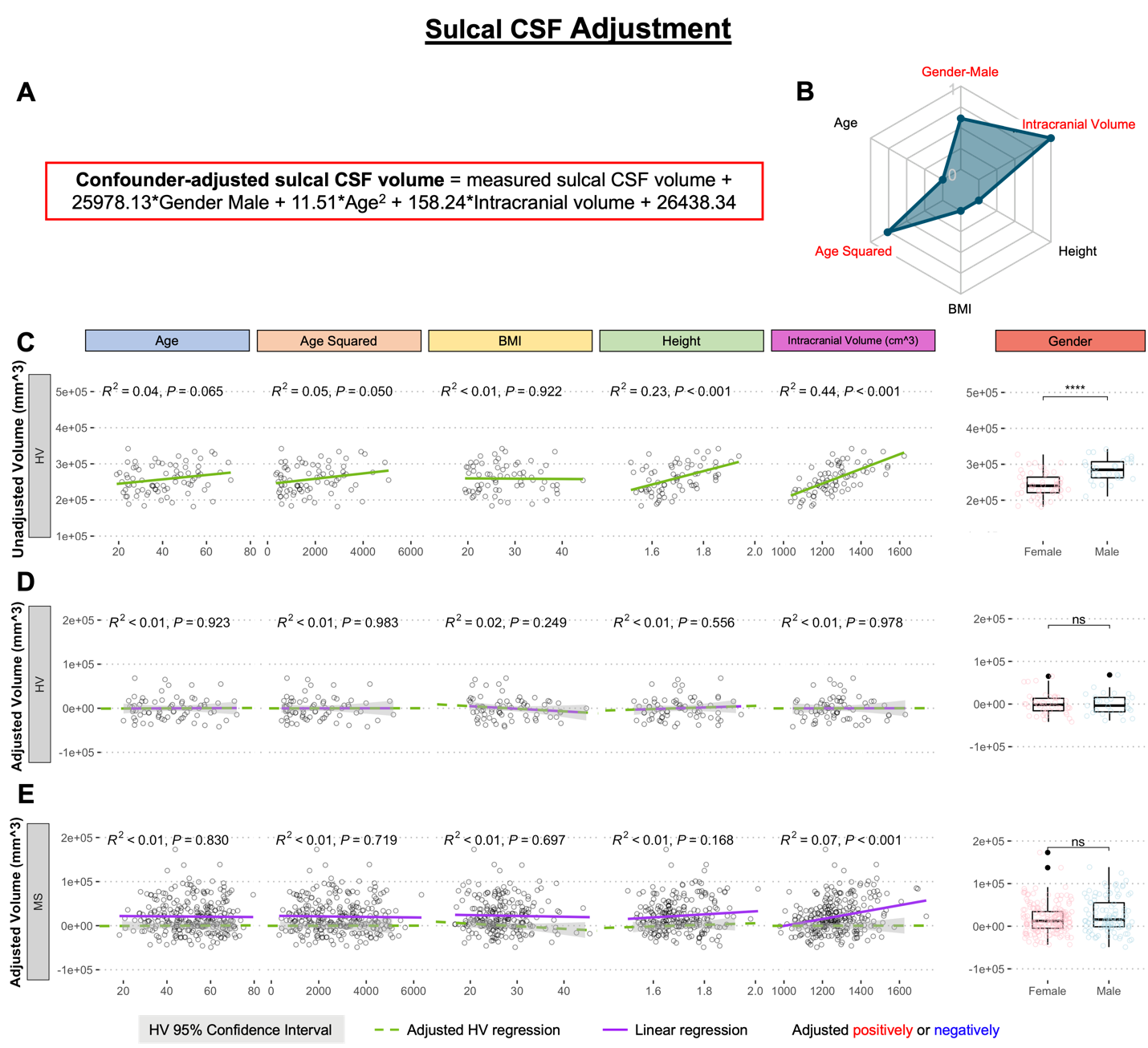

**Supplementary Figure S14. Adjustment of single MRI biomarker (i.e., sulcal CSF volume) for six physiological confounders. A.** The final equation for the confounder-adjusted sulcal CSF volume is shown at the top in red outline. This equation was derived from multiple linear regression models as described in the Method section. **B.** In the top right corner is resulting radar chart that show proportional weights of the applied confounder adjustment, with confounders with lowest weights (i.e., the innermost circle) representing zeros. **C.** Univariate linear regression models between each tested confounder on x-axis (first 5 graphs) or gender (sixth graph) and measured sulcal CSF volume on the y-axis in 80 healthy volunteers (HV). **D.** Same univariate regressions in the HV cohort after applying adjustment formula show no remaining effect of confounders. **E.** Applying HV-derived adjustment formula to MS cohort shows residual effect of intracranial volume.

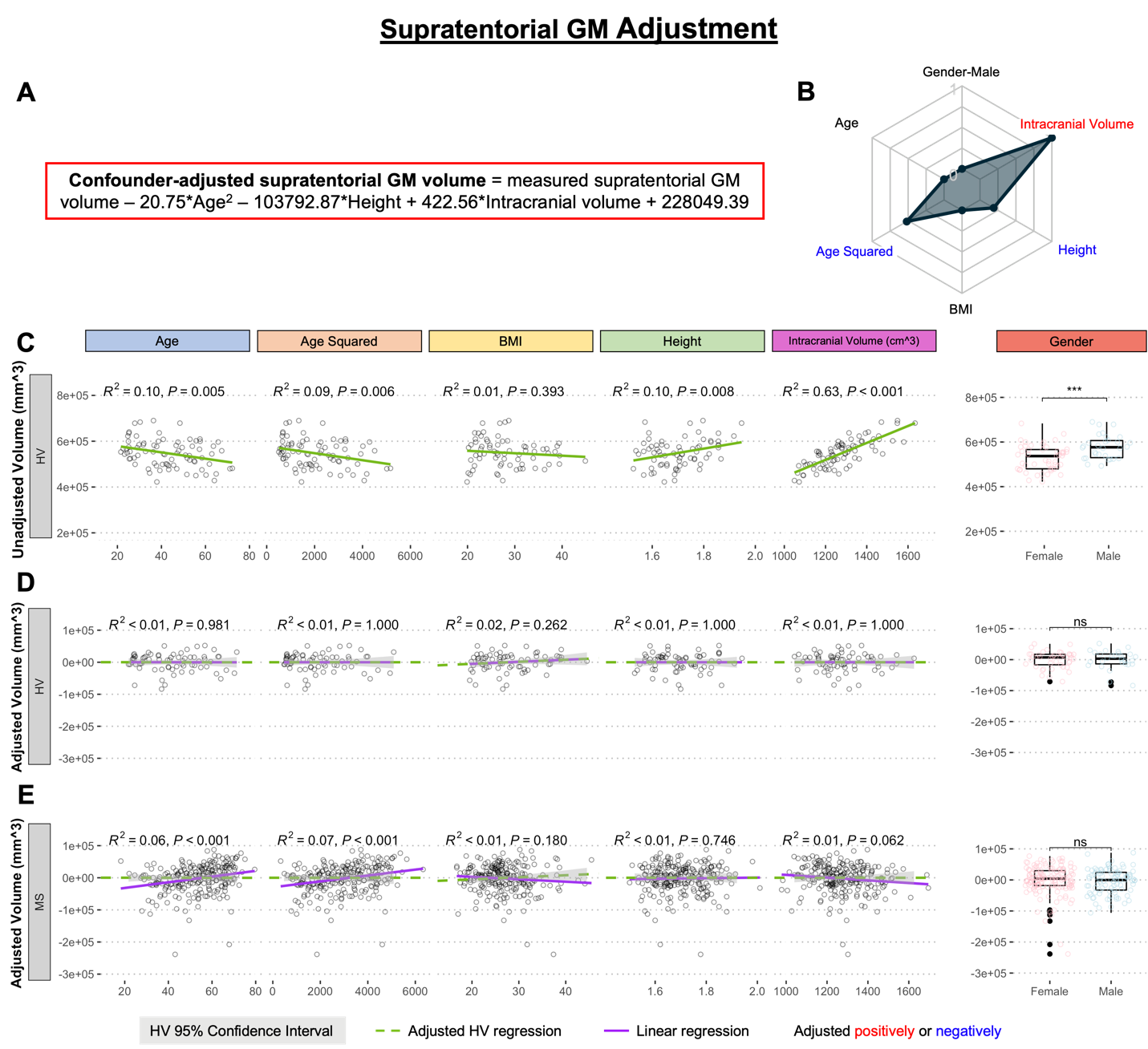

**Supplementary Figure S15. Adjustment of single MRI biomarker (i.e., supratentorial GM volume) for six physiological confounders. A.** The final equation for the confounder-adjusted supratentorial GM volume is shown at the top in red outline. This equation was derived from multiple linear regression models as described in the Method section. **B.** In the top right corner is resulting radar chart that show proportional weights of the applied confounder adjustment, with confounders with lowest weights (i.e., the innermost circle) representing zeros. **C.** Univariate linear regression models between each tested confounder on x-axis (first 5 graphs) or gender (sixth graph) and measured supratentorial GM volume on the y-axis in 80 healthy volunteers (HV). **D.** Same univariate regressions in the HV cohort after applying adjustment formula show no remaining effect of confounders. **E.** Applying HV-derived adjustment formula to MS cohort shows residual effect of age and age^2.

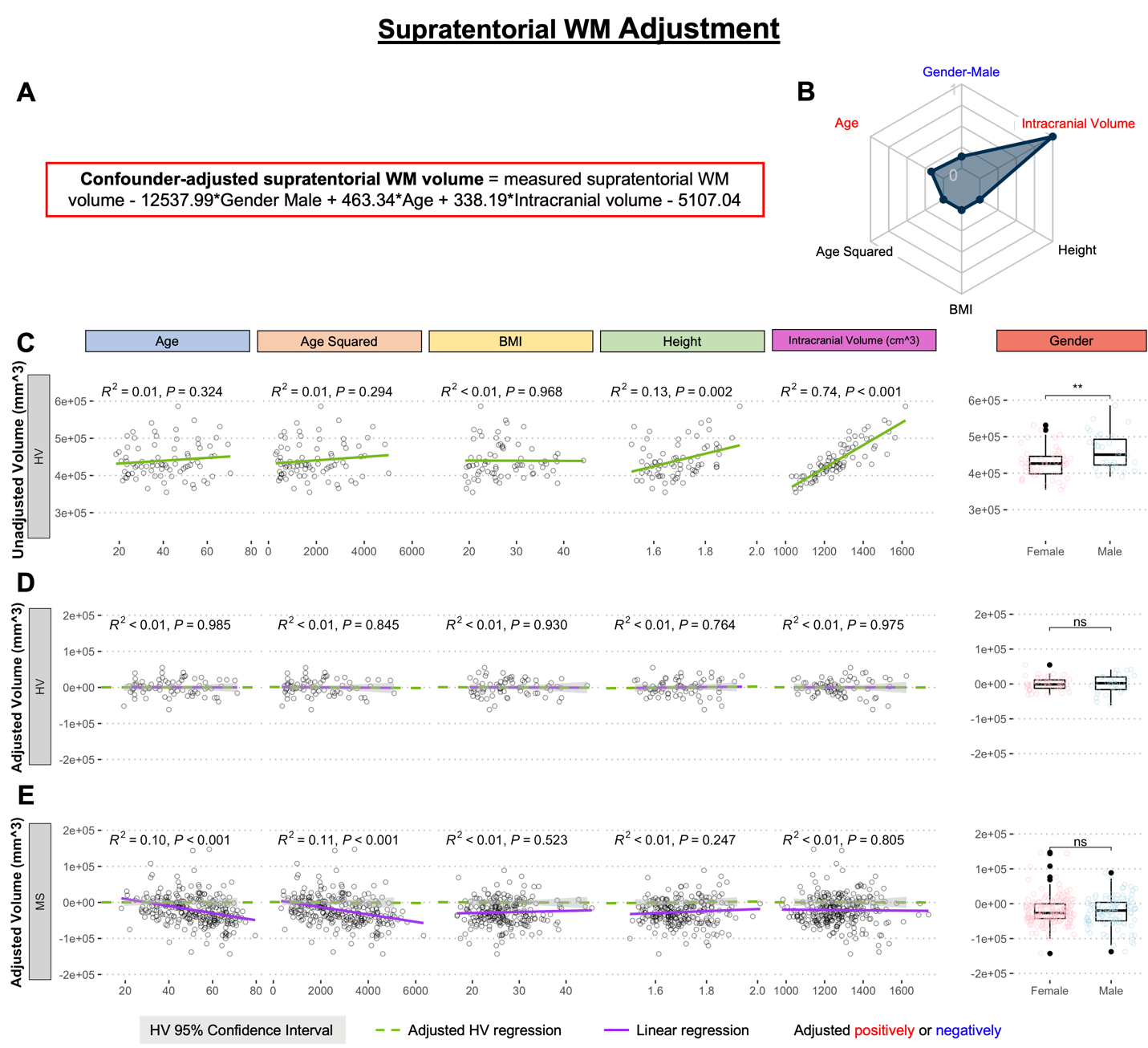

**Supplementary Figure S16. Adjustment of single MRI biomarker (i.e., supratentorial WM volume) for six physiological confounders. A.** The final equation for the confounder-adjusted supratentorial WM volume is shown at the top in red outline. This equation was derived from multiple linear regression models as described in the Method section. **B.** In the top right corner is resulting radar chart that show proportional weights of the applied confounder adjustment, with confounders with lowest weights (i.e., the innermost circle) representing zeros. **C.** Univariate linear regression models between each tested confounder on x-axis (first 5 graphs) or gender (sixth graph) and measured supratentorial WM volume on the y-axis in 80 healthy volunteers (HV). **D.** Same univariate regressions in the HV cohort after applying adjustment formula show no remaining residual effect of confounders. **E.** Applying HV-derived adjustment formula to MS cohort shows residual effect of age and age^2.

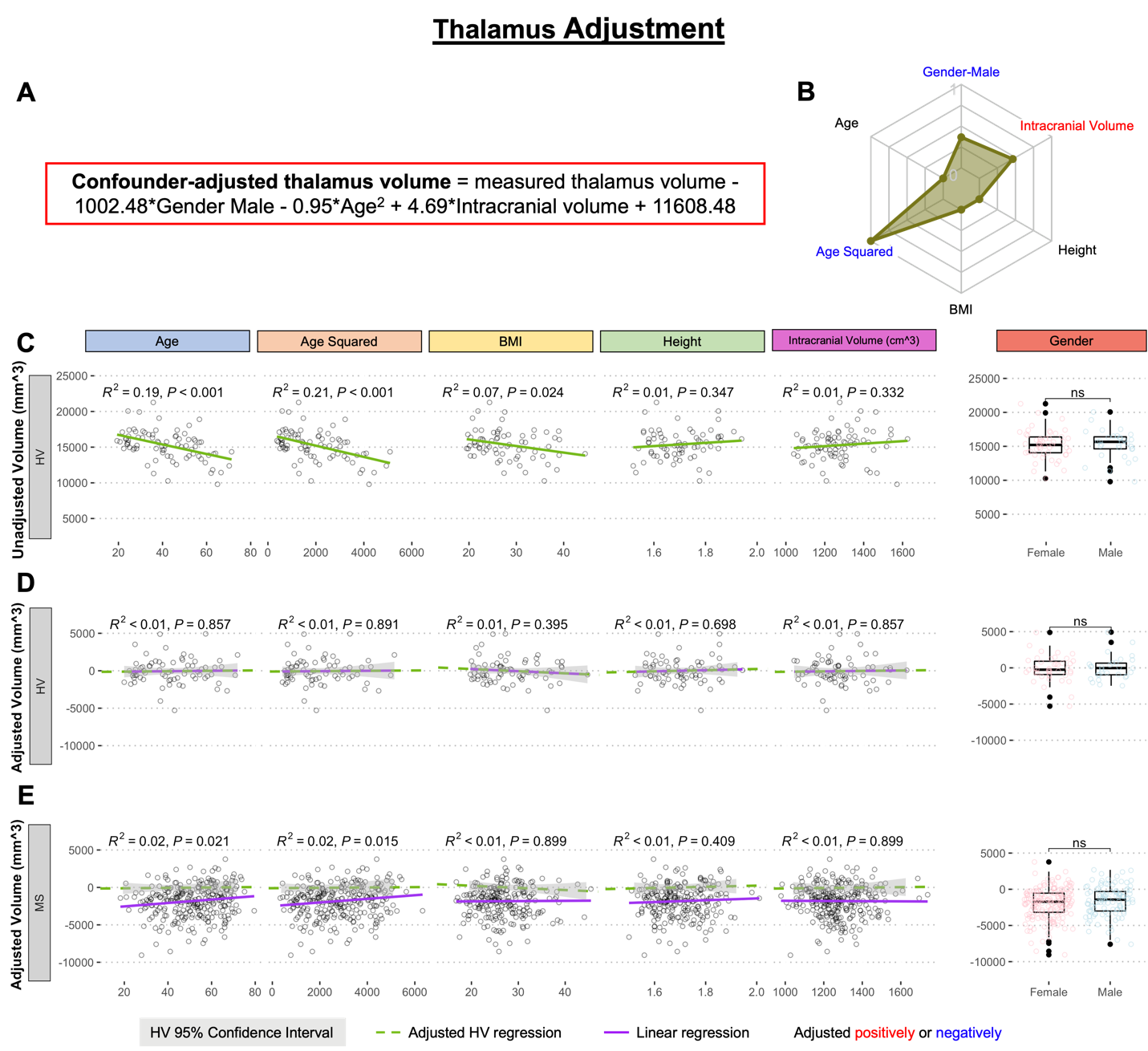

**Supplementary Figure S17. Adjustment of single MRI biomarker (i.e., thalamus volume) for six physiological confounders. A.** The final equation for the confounder-adjusted thalamus volume is shown at the top in red outline. This equation was derived from multiple linear regression models as described in the Method section. **B.** In the top right corner is resulting radar chart that show proportional weights of the applied confounder adjustment, with confounders with lowest weights (i.e., the innermost circle) representing zeros. **C.** Univariate linear regression models between each tested confounder on x-axis (first 5 graphs) or gender (sixth graph) and measured thalamus volume on the y-axis in 80 healthy volunteers (HV). **D.** Same univariate regressions in the HV cohort after applying adjustment formula show no remaining residual effect of confounders. **E.** Applying HV-derived adjustment formula to MS cohort shows no residual effect, when considering Bonferroni adjustment for multiple comparisons (i.e., p<0.05/12 = p<0.004).

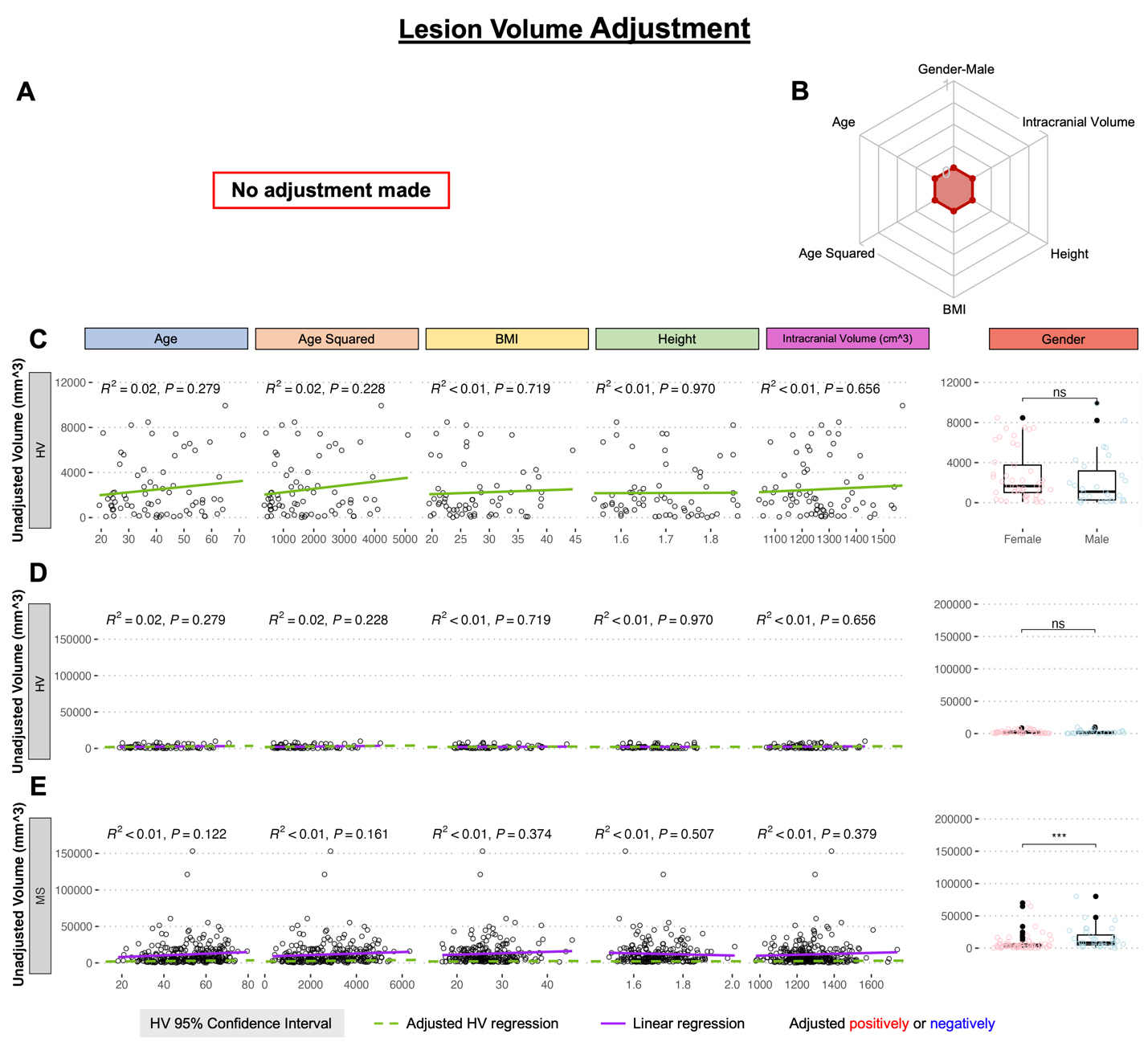

**Supplementary Figure S18. Adjustment of single MRI biomarker (i.e., lesion volume) for six physiological confounders. A.** No confounder-adjustment was made for lesion volume. **B.** In the top right corner is resulting radar chart that show proportional weights of the applied confounder adjustment, with confounders with lowest weights (i.e., the innermost circle) representing zeros. **C.** Univariate linear regression models between each tested confounder on x-axis (first 5 graphs) or gender (sixth graph) and measured lesion volume on the y-axis in 80 healthy volunteers (HV). **D.** No univariate linear regression of confounders was seen in the HV cohort. **E.** No univariate linear regression of confounders was seen in the MS cohort.

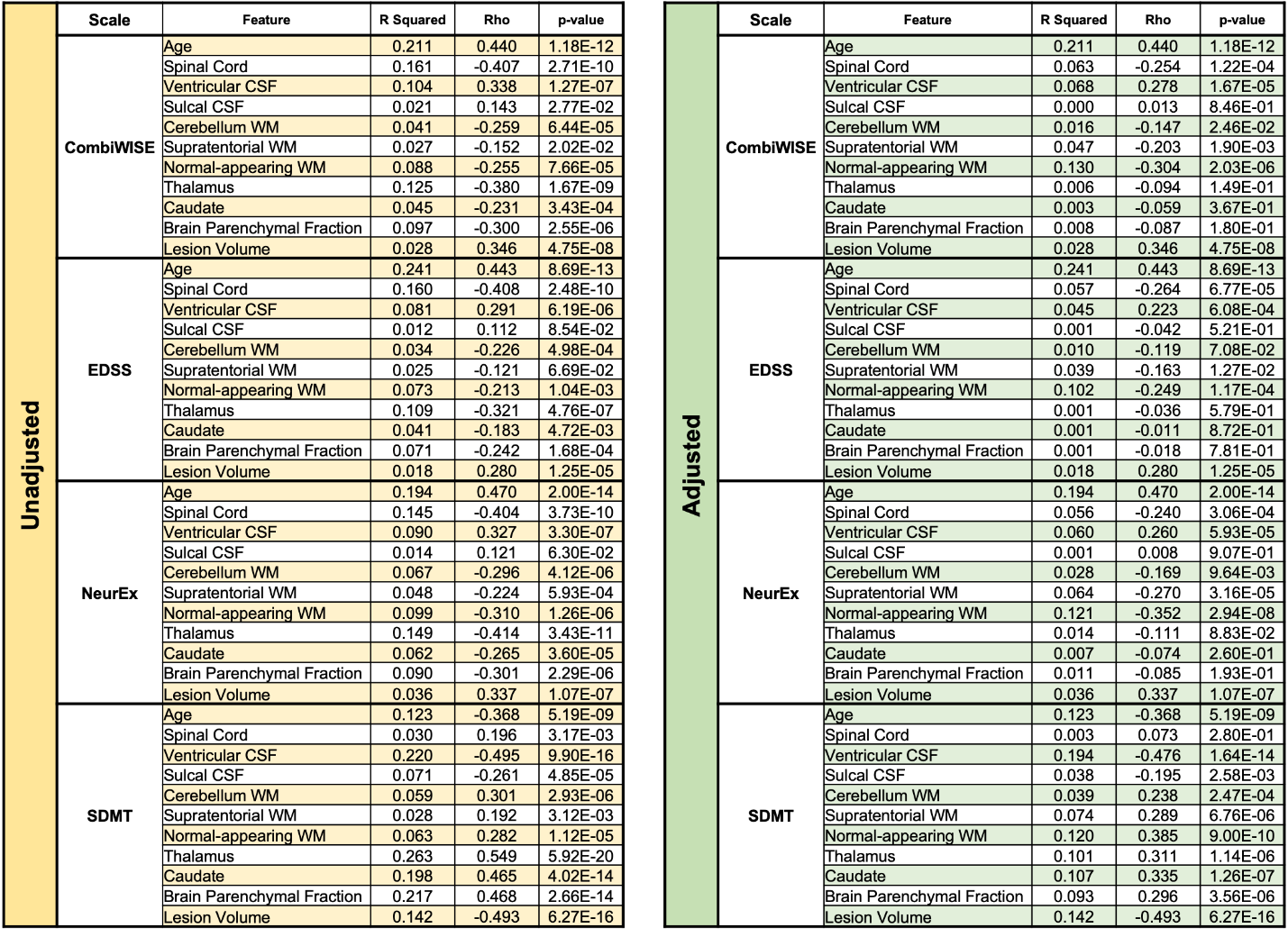

**Supplementary Table S1. Unilateral correlations of disability outcomes with MRI features.**

The left panel shows R squared, Spearman correlation coefficients (Rho), and p-values between unadjusted MRI features and disability outcomes. The right panel shows R squared, Spearman correlation coefficients, and p-values between confounder-adjusted MRI features and disability outcomes.

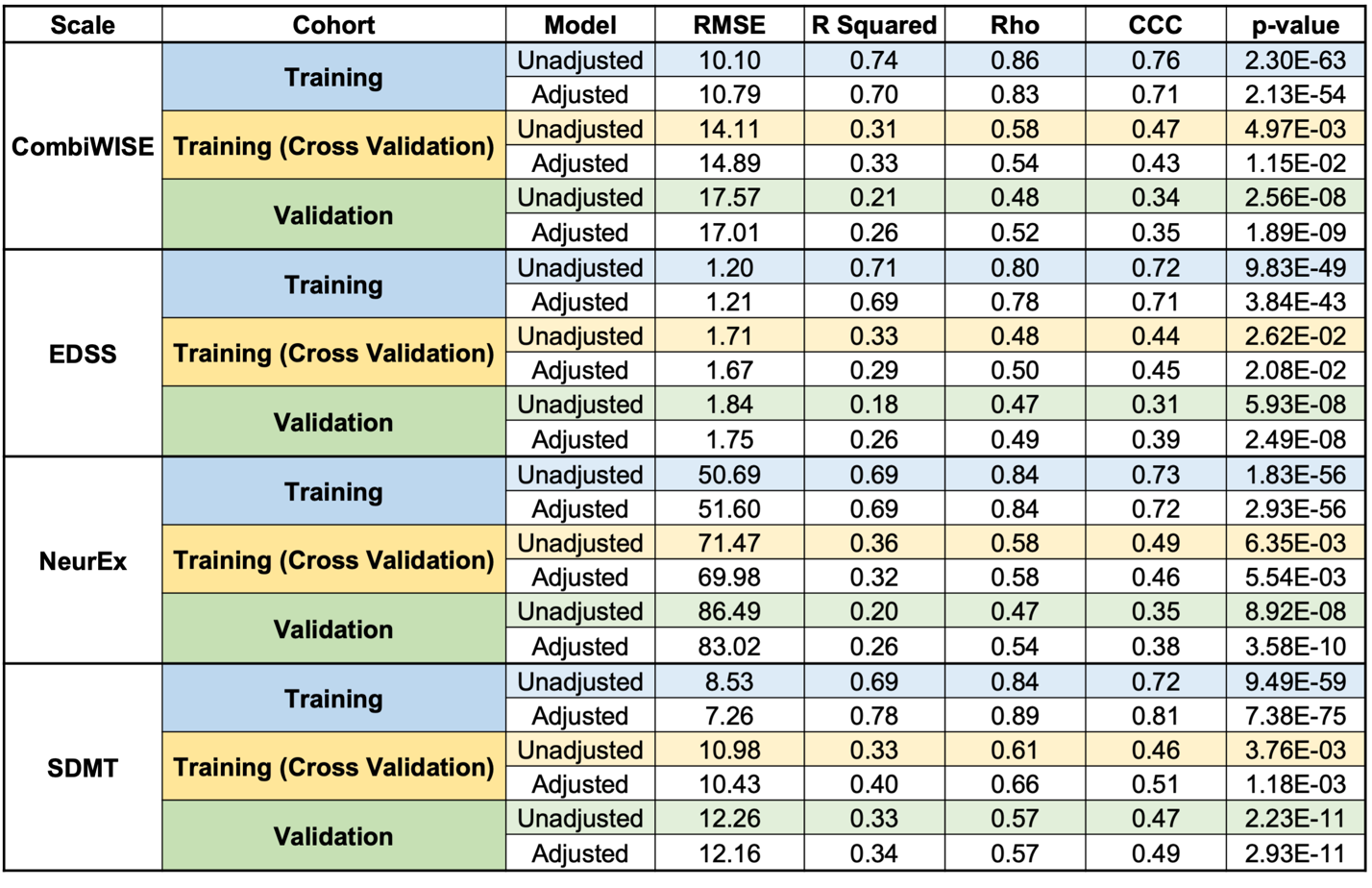

**Supplementary Table S2. Statistics on model performances in each cohort.**

This table shows model performances in predicting disability scales individually within the training cohort, training cohort with cross validation, and independent validation cohort.
